# Para-infectious brain injury in COVID-19 persists at follow-up despite attenuated cytokine and autoantibody responses

**DOI:** 10.1101/2023.04.03.23287902

**Authors:** Benedict D. Michael, Cordelia Dunai, Edward J. Needham, Kukatharmini Tharmaratnam, Robyn Williams, Yun Huang, Sarah A. Boardman, Jordan Clark, Parul Sharma, Krishanthi Subramaniam, Greta K. Wood, Ceryce Collie, Richard Digby, Alexander Ren, Emma Norton, Maya Leibowitz, Soraya Ebrahimi, Andrew Fower, Hannah Fox, Esteban Tato, Mark Ellul, Geraint Sunderland, Marie Held, Claire Hetherington, Franklyn Nkongho, Alish Palmos, Alexander Grundmann, James P. Stewart, Michael Griffiths, Tom Solomon, Gerome Breen, Alasdair Coles, Jonathan Cavanagh, Sarosh R. Irani, Angela Vincent, Leonie Taams, David K. Menon

**Affiliations:** University of Liverpool; NIHR Health Protection Research Unit (HPRU) in Emerging and Zoonotic Infections at University of Liverpool; University of Cambridge; University of Oxford; King’s College London; University of Southampton; University of Glasgow

## Abstract

We measured brain injury markers, inflammatory mediators, and autoantibodies in 203 participants with COVID-19; 111 provided acute sera (1-11 days post admission) and 56 with COVID-19-associated neurological diagnoses provided subacute/convalescent sera (6-76 weeks post-admission). Compared to 60 controls, brain injury biomarkers (Tau, GFAP, NfL, UCH-L1) were increased in acute sera, significantly more so for NfL and UCH-L1, in patients with altered consciousness. Tau and NfL remained elevated in convalescent sera, particularly following cerebrovascular and neuroinflammatory disorders. Acutely, inflammatory mediators (including IL-6, IL-12p40, HGF, M-CSF, CCL2, and IL-1RA) were higher in participants with altered consciousness, and correlated with brain injury biomarker levels. Inflammatory mediators were lower than acute levels in convalescent sera, but levels of CCL2, CCL7, IL-1RA, IL-2Rα, M-CSF, SCF, IL-16 and IL-18 in individual participants correlated with Tau levels even at this late time point. When compared to acute COVID-19 patients with a normal GCS, network analysis showed significantly altered immune responses in patients with acute alteration of consciousness, and in convalescent patients who had suffered an acute neurological complication. The frequency and range of autoantibodies did not associate with neurological disorders. However, autoantibodies against specific antigens were more frequent in patients with altered consciousness in the acute phase (including MYL7, UCH-L1, GRIN3B, and DDR2), and in patients with neurological complications in the convalescent phase (including MYL7, GNRHR, and HLA antigens). In a novel low-inoculum mouse model of SARS-CoV-2, while viral replication was only consistently seen in mouse lungs, inflammatory responses were seen in both brain and lungs, with significant increases in CCL4, IFNγ, IL-17A, and microglial reactivity in the brain. Neurological injury is common in the acute phase and persists late after COVID-19, and may be driven by a para-infectious process involving a dysregulated host response.

**Graphical abstract:** (a) The acute cohort (days 1-11 post-hospitilisation) showed elevated pro-inflammatory cytokines, brain injury markers, and autoantibodies. The sub-acute/convalescent cohort (weeks to months post-COVID+ve test) retained elevated brain injury markers but lower proinflammatory cytokines and autoantibodies.
(b) The mouse model of para-infectious brain with no active viral replication, had increased cytokines (IFNγ and IL-17A) and microglia reactivity (increased Iba1 expression).
Created using Biorender.

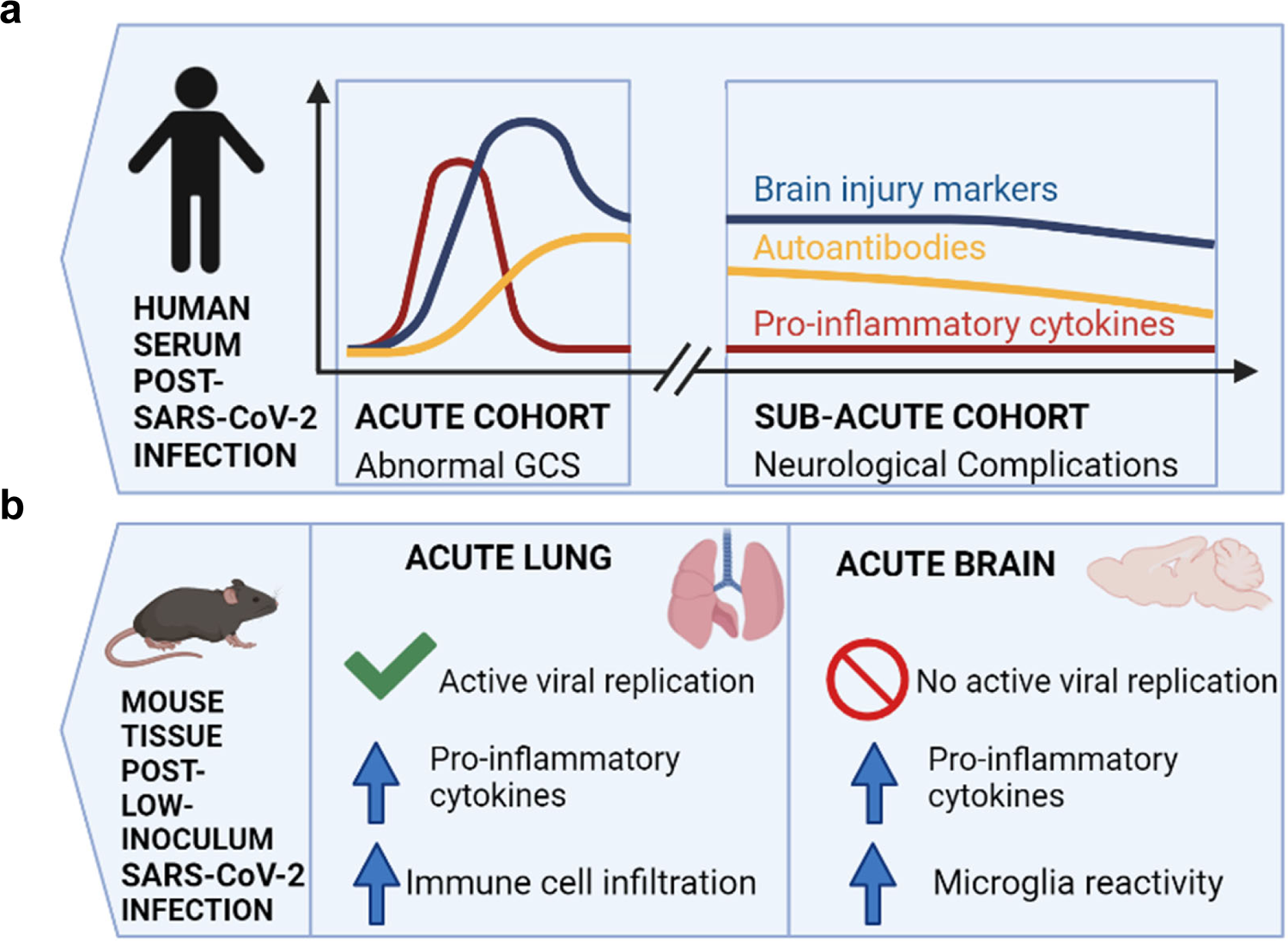

## Introduction

At the beginning of the COVID-19 pandemic, neurological complications occurred in approximately one-third of hospitalised patients^1^ and even in those with mild COVID-19 infection^2^. Whilst these neurological ‘complications’ were often mild (headache and myalgia), it became clear that more significant neurological sequelae were observed, including encephalitis/encephalopathies, Guillain Barre Syndrome, and stroke^3–5^.

Although i*n vitro* studies show that SARS-CoV-2 can infect neurons and astrocytes^6, 7^, autopsy studies indicate that direct viral invasion is unlikely to be a cause of neurological dysfunction *in vivo*. Post-mortem studies failed to detect viral infection by immunohistochemistry in the majority of cases, and viral qPCR levels were often low and may simply have reflected viraemia^8–10^. In addition, virus and/or anti-viral antibodies were rarely found in cerebrospinal fluid (CSF)^11^. Thus, it seems more likely that the virus affects the brain indirectly. This could be through peripherally generated inflammatory mediators, immune cells, autoantibodies and/or blood brain barrier changes associated with endothelial damage^12, 13^. Immune infiltrates have been found in autopsy studies, including neutrophils and T cells, although agonal effects could not be excluded^14^. Elevated IL-6 and D-dimer levels at admission are associated with neurological complications, including thrombosis, stroke, cognitive and memory deficits, regardless of respiratory disease severity^15–17^. The brain injury markers NfL and GFAP, and inflammatory cytokines are elevated in COVID-19 and scale with severity, but their relationship to neuropathology has not been reported^18–21^. Finally, specific neuronal autoantibodies have been reported in some neurological patients raising the possibility of para- or post-infectious autoimmunity^12, 22^.

To assess the relationship between host response and biomarkers of neurological injury, we studied two large, multisite cohorts which, in combination, provided acute, subacute and convalescent sera from COVID-19-positive (COVID+ve) participants. We measured brain injury markers, a range of cytokines and associated inflammatory mediators, and autoantibodies, in these samples, and related them to reduced levels of consciousness (defined as a Glasgow Coma Scale Score [GCS] GCS≤14) in the acute phase, or the history of a neurological complication of COVID-19 in convalescent participants. We further explored these relationships between host response and brain injury in a novel mouse model of low-inoculum intranasal SARS-CoV-2 infection.

## Results

### COVID-19 results in acute elevation of serum brain injury biomarkers, more so in participants with abnormal Glasgow coma scale (GCS) score

We used sera from the ISARIC study, obtained 1-11 days post admission, including 111 participants with COVID-19 of varying severity and 60 COVID-19-ve healthy controls (labelled Control). Participants were stratified by normal (n = 76) or abnormal (n = 35) Glasgow coma scale (labelled GCS=15 or GCS≤14, respectively) scores to provide a proxy for neurological dysfunction (Fig. 1a). GFAP (glial fibrillary acidic protein, marker of astrocyte injury), UCH-L1 (a marker of neuronal cell body injury), and NfL (neurofilament light) and Tau (both markers of axonal and dendritic injury) were measured. Overall, serum levels of NfL, GFAP, and Tau were significantly higher in COVID-19 participants compared to the COVID-19–ve healthy controls, but as shown in Fig. 1b-e, those participants with abnormal GCS scores had higher levels of NfL and UCH-L1 than those with normal GCS scores. Thus, all four biomarkers were raised in COVID-19 participants (both GCS=15 and GCS≤14) but, in addition, axonal and neuronal body injury biomarkers discriminated between participants with and without reduced GCSs.

**Figure 1:**
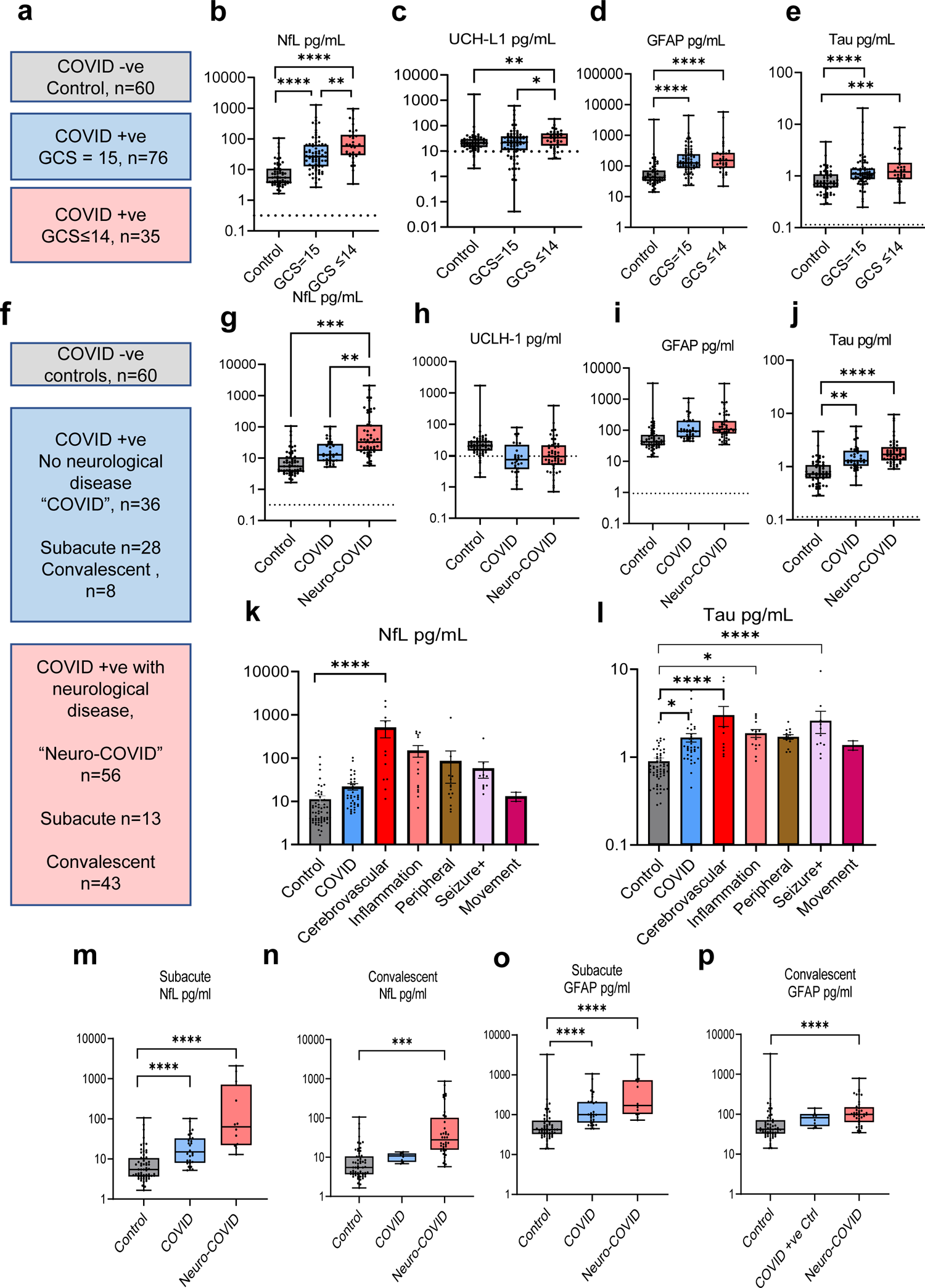
Brain injury markers NfL and UCH-L1 are elevated acutely in COVID-19 patients with an abnormal Glasgow coma scale score (GCS) and correlate with cytokine/chemokine networks. **(a)** Day 1-11 admission acute serum brain injury markers were assessed by Simoa: **(b)** NfL, **(c)** UCH-L1, **(d)** GFAP, and **(e)**Tau. All four were elevated in COVID-19 cases with normal Glasgow coma scale scores (GCS) relative to controls overall. NfL and UCH-L1 were further elevated in patients with an abnormal GCS. Dotted lines show lower limit of quantification. **(f-j)** Brain injury markers were measured by Simoa in serum from the sub-acute (<6 weeks from positive SARS-CoV-2 test) and convalescent phase (>6 weeks). **(k)** Within the sub-acute and convalescent COVID-19 neurological cases, the highest levels of NfL were observed in patients who had suffered a cerebrovascular at the time of SARS-CoV-2 infection. **(l)** Within the neurological cases, Tau levels in the cerebrovascular, CNS inflammatory and seizure conditions were significantly higher compared to healthy controls. **(m,n)** Serum NfL remained elevated in both the sub-acute and convalescent phases in cases with an acute neurological complication of COVID-19 as opposed to COVID-19 controls. **(o,p)** GFAP was elevated in neurological cases in the sub-acute but not convalescent phase. *Group comparisons are by Kruskal-Wallis test, pairwise comparisons by Mann-Whitney U test, Volcano plots use multiple Mann-Whitney U tests with a false discovery rate set to 1%, and correlations are Pearson’s coefficients (* p<0.05, ** p<0.01, *** p<0.001, **** p<0.0001)*.

### Brain injury biomarkers remain elevated in the subacute and convalescent phase in participants who have had a CNS complication of COVID-19

To ask whether these findings persisted in participants recovering from COVID-19-related neurological complications, ninety-two COVID-19+ve subjects were recruited to the COVID-Clinical Neuroscience Study (COVID-CNS), 56 who had had a new neurological diagnosis that developed as an acute complication of COVID-19 (group labelled “neuro-COVID”), and 36 with no such neurological complication (group labelled “COVID”, Fig. 1f, Table 1, Extended Data Table 1 and 2). When compared to the same healthy controls (n = 60), across all time-points, both COVID-19+ve subgroups (COVID and neuro-COVID) showed increased levels of NfL and Tau (but not UCH-L1 or GFAP (Fig. 1g-j, Extended Data Table 1). Furthermore, participants recovering from neuro-COVID had significantly higher levels of NfL, and a trend towards higher levels of Tau, than the COVID participants (Fig. 1g,j). Highest NfL serum levels were present in participants with cerebrovascular conditions, whereas Tau was elevated in participants with cerebrovascular, CNS inflammation and peripheral nerve complications (Fig. 1k,l). We then separately compared the two cohorts at subacute and convalescent follow up periods (less than and over six weeks respectively). NfL and GFAP levels remained elevated in all COVID-19 participants in the subacute period, but only remained elevated beyond 6 weeks in participants who had suffered an acute neurological complication (neuro-COVID, Fig. 1 m-p; Extended Data Fig. 1a). The presence of elevated brain injury biomarkers in the acute phase of COVID-19 confirms previous findings,^12^ but the elevated levels of NfL and GFAP in those who had acute neurological complications suggests ongoing neuroglial injury, months after the acute illness.

**Table 1:**
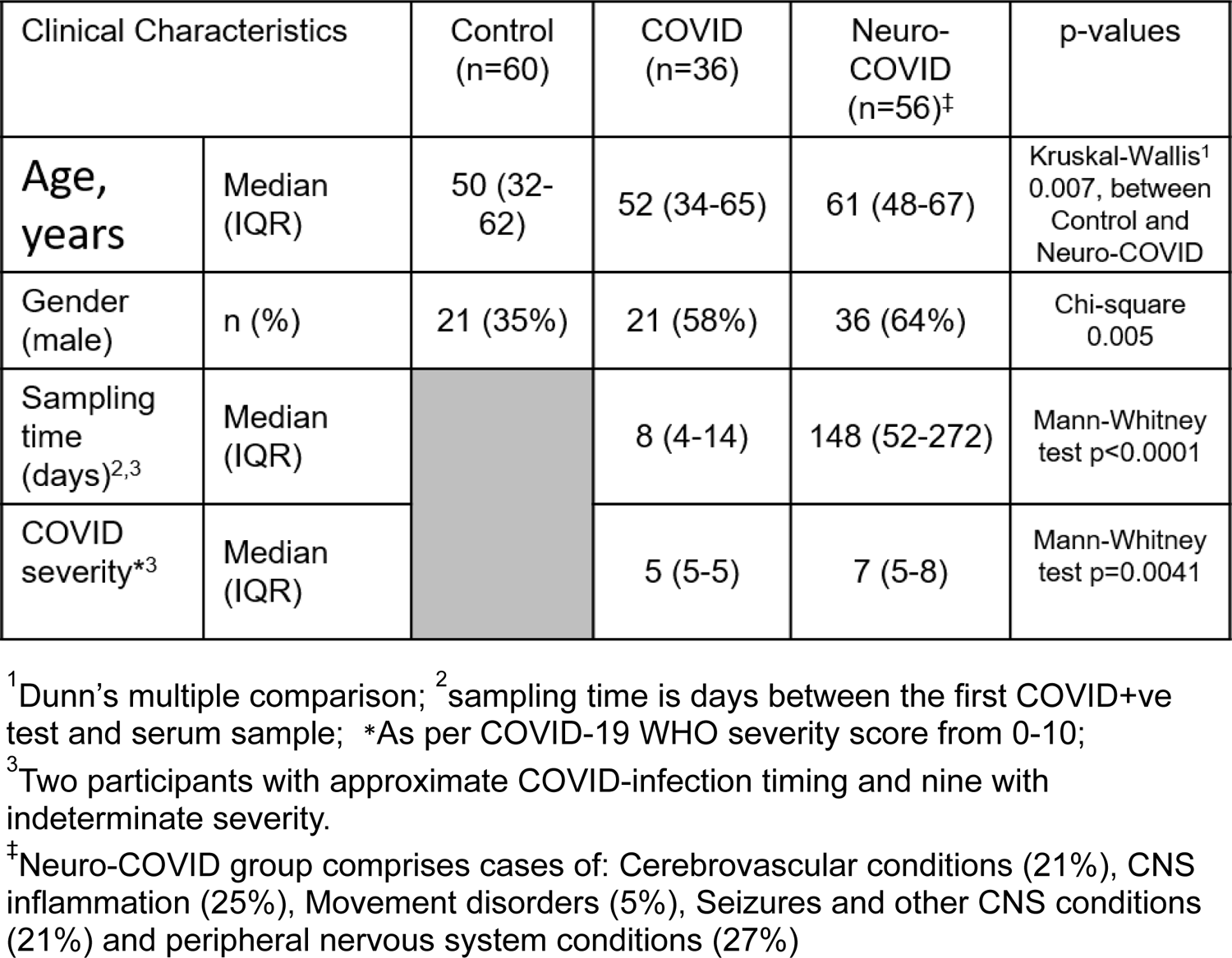
Clinical characteristics of healthy controls and COVID-CNS Participants

| Clinical Characteristics |  | Control<br>(n=60) | COVID<br>(n=36) | Neuro-COVID<br>(n=56)<sup>‡</sup> | p-values |
| --- | --- | --- | --- | --- | --- |
| **Age,**<br>**years** | Median<br>(IQR) | 50 (32-62) | 52 (34-65) | 61 (48-67) | Kruskal-Wallis<sup>1</sup><br>0.007, between<br>Control and<br>Neuro-COVID |
| Gender<br>(male) | n (%) | 21 (35%) | 21 (58%) | 36 (64%) | Chi-square<br>0.005 |
| Sampling<br>time<br>(days)<sup>2,3</sup> | Median<br>(IQR) |  | 8 (4-14) | 148 (52-272) | Mann-Whitney<br>test p<0.0001 |
| COVID<br>severity<sup>*3</sup> | Median<br>(IQR) |  | 5 (5-5) | 7 (5-8) | Mann-Whitney<br>test p=0.0041 |

<sup>1</sup>Dunn's multiple comparison; <sup>2</sup>sampling time is days between the first COVID+ve test and serum sample; <sup>\*</sup>As per COVID-19 WHO severity score from 0-10;
<sup>3</sup>Two participants with approximate COVID-infection timing and nine with indeterminate severity.
<sup>‡</sup>Neuro-COVID group comprises cases of: Cerebrovascular conditions (21%), CNS inflammation (25%), Movement disorders (5%), Seizures and other CNS conditions (21%) and peripheral nervous system conditions (27%)

### Clinical and biomarker evidence of neurological insult levels are associated with levels of innate inflammatory mediators in the acute phase of COVID-19

To explore whether the acute and persistent elevation of brain injury biomarkers we observed in participants with COVID-19 was associated with an acute inflammatory response, we measured a panel of 48 inflammatory mediators in serum at the same time points. In the ISARIC samples, six mediators were significantly higher in participants with an abnormal GCS than in those with a normal GCS (interleukin [IL]-6, hepatocyte growth factor [HGF], IL-12p40, IL-1RA, CCL2 and macrophage colony stimulating factor [M-CSF]), indicating increased innate inflammation (Fig. 2a, Extended Data Fig. 2a). Pearson’s correlation tests identified correlations between these significant biomarkers in an interrelated pro-inflammatory network (Fig. 2b,c), and unsupervised Euclidean hierarchical cluster analysis revealed clusters of pro-inflammatory mediators elevated together (Fig. 2d). The first cluster incorporated the IL-1 family (including IL-1RA), interferons and M-CSF, and the second cluster included IL-6, CCL2, CXCL9, HGF, and IL-12p40 (boxes in Fig. 2d). Brain injury biomarkers correlated with elevations in these inflammatory mediators: GFAP and UCLH-1 correlated with a number of mediators, while Tau and NfL correlated strongly with HGF and IL-12p40 in the second cluster (Extended Data Table 3).

**Figure 2:**
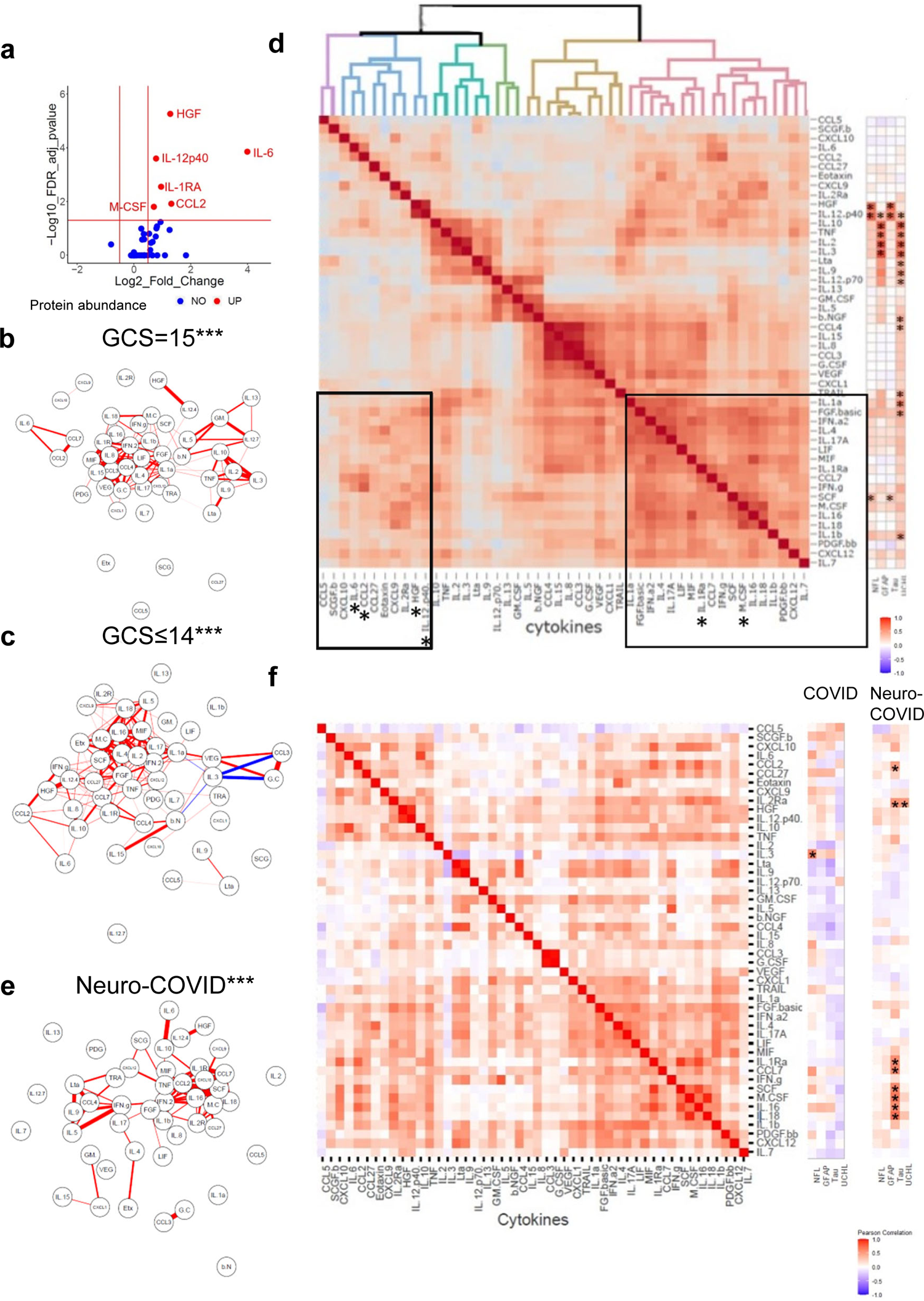
In the sub-acute/convalescent phase NfL remains elevated in those who have suffered a CNS complication, especially stroke and CNS inflammation, despite down-regulation of pro-inflammatory mediators. Serum mediators from ISARIC and COVID-CNS were assessed by Luminex. **(a)** A volcano plot was generated to identify those mediators elevated in patients with an abnormal GCS and **(b,c)** a network analysis identified the highest correlations between the mediators (*significantly different from ISARIC GCS=15 by Steiger test p<0.001) and **(d)** unbiased Euclidean hierarchical cluster and correlation analyses conducted, identifying two clusters of up-regulation of several pro-inflammatory mediators in concert. The first group included interleukin (IL)-6, IL-12p40, CCL2, CXCL9 and hepatocyte growth factor (HGF) and the second group included the IL-1 family, interferons, and macrophage colony stimulating factor (M-CSF). The heat map on the right shows cytokine correlations with brain injury biomarkers, highlighting those mediators that were significantly correlated with asterisks. **(e)** Network analysis and heatmaps of correlations between mediators did not demonstrate the tight interconnectedness which had been identified in acute samples (*significantly different from ISARIC GCS=15 and GCS≤14 by Steiger test p<0.001) and **(f)** several mediators significantly correlated with Tau. *Group comparisons are by Kruskal-Wallis test, pairwise comparisons by Mann-Whitney U test, Volcano plots use multiple Mann-Whitney U tests with a false discovery rate set to 1%, and correlations are Pearson’s coefficients (* p<0.05, ** p<0.01, *** p<0.001, **** p<0.0001)*.

A more stringent analysis of median-centred cytokine data (which corrected for within-participant skewing of mediator levels) confirmed that HGF and IL-12p40 were higher in the abnormal GCS COVID-19 participants, and correlated with cognate NfL levels (Extended Data Table 4). Taken together these data suggest that activation of the innate immune system was related to both clinical and blood biomarker evidence of CNS insult.

### Inflammatory mediators are not elevated across the participant cohort at late timepoints after COVID-19; but late Tau elevations correlate with levels of several inflammatory mediators

In contrast to the acute data, the levels of cytokines and associated mediators were lower when measured during the subacute and convalescent periods even in those who had suffered neurological complications of COVID-19 (group labelled “neuro-COVID”. Extended Data Fig.2b). Indeed IL-18 was downregulated in the neuro-COVID cases compared to controls (Extended Data Table 1, Extended Data Fig. 2c; also with median-centred data analysis), and the correlations between cytokines and associated mediators no longer displayed the same tight clusters (Fig. 2e,f). GFAP remained elevated during the convalescent phase of neurological complications (Fig. 1p) but did not show correlations with the inflammatory mediators. Similarly NfL was higher overall in those with neurological complications (Fig. 1n) but there were no significant correlations with inflammatory mediators (Fig. 2f). However, Tau remained elevated overall in those with neurological complications ((1.7 (1.3, 2.2) pg/mL versus 1.3 (1.1, 1.9) pg/mL)) and levels correlated with eight immune mediators including CCL2, IL-1RA, IL-2Rα and M-CSF along with CCL7, stem cell factor (SCF), IL-16 and IL-18 (Fig. 2f, Extended Data Table 5, Extended Data Fig. 2d). This last association was specific to the late phase of the illness and was not found in acute COVID-19.

### Cytokine networks are significantly altered in patients with neurological complications of COVID-19: both acute encephalopathy, and those recovering from a neurological complication

We used graph theoretical approaches to characterise cytokine networks in our three groups of patients: acute COVID-19 with a normal GCS; acute COVID-19 with altered consciousness (GCS≤14), and convalescent patients recovering from a neurological complication of COVID-19 (neuro-COVID). Participants with both acute and chronic neurological consequences of COVID-19 (GCS≤14) and Neuro-COVID both showed cytokine networks that were different from COVID-19 patients with no neurological problems (Fig. 2 b,c,e; p<0.001, Steiger test), suggesting a specific dysregulated innate immune response that is associated with both acute and chronic neurological complications of the illness.

### COVID-19 is associated with an acute polyclonal inflammatory response overall and with autoantibodies to viral antigen and some CNS autoantigens in those with an abnormal GCS score

Given past reports of autoantibody responses following COVID-19^12, 22^, we sought evidence of similar dysregulated adaptive immune responses in our participant cohorts. We used a bespoke protein chip array of 153 viral and tissue proteins to measure IgM and IgG reactivity in the acute phase ISARIC sera. The data from individual participants were normalized and the median fluorescence intensity per participant and Z-scores of each participant were compared to healthy control data, to determine positive reactivity to the different antigens (with a threshold for detection set at three standard deviations above COVID-ve controls for each antigen; see Extended Data Table 6, Extended Data Fig.3a, 4a). IgM and IgG responses in COVID+ve participants showed greater reactivity overall (both GCS=15 and GCS≤14), compared to the controls, with no difference in normalised fluorescence Z scores or the number of participants with IgG ‘hits’ (a Z-score >3) between those with normal or abnormal GCS score (Fig. 3a,b, Extended Data Fig. 3a). However, several IgM and IgG autoantibodies, including against the CNS antigens UCH-L1, GRIN3B and DRD2, along with the cardiac antigen, myosin light chain (MYL)-7, were present in a greater proportion of participants with an abnormal GCS score, as were antibodies to spike protein (Fig. 3c, Extended Data Fig. 3b). None of the antibodies correlated significantly with levels of brain injury markers (Extended Data Fig. 3c), but they did show correlations with each other (Fig. 3d), suggesting a non-specific antibody response in some individuals during the acute phase.

**Figure 3:**
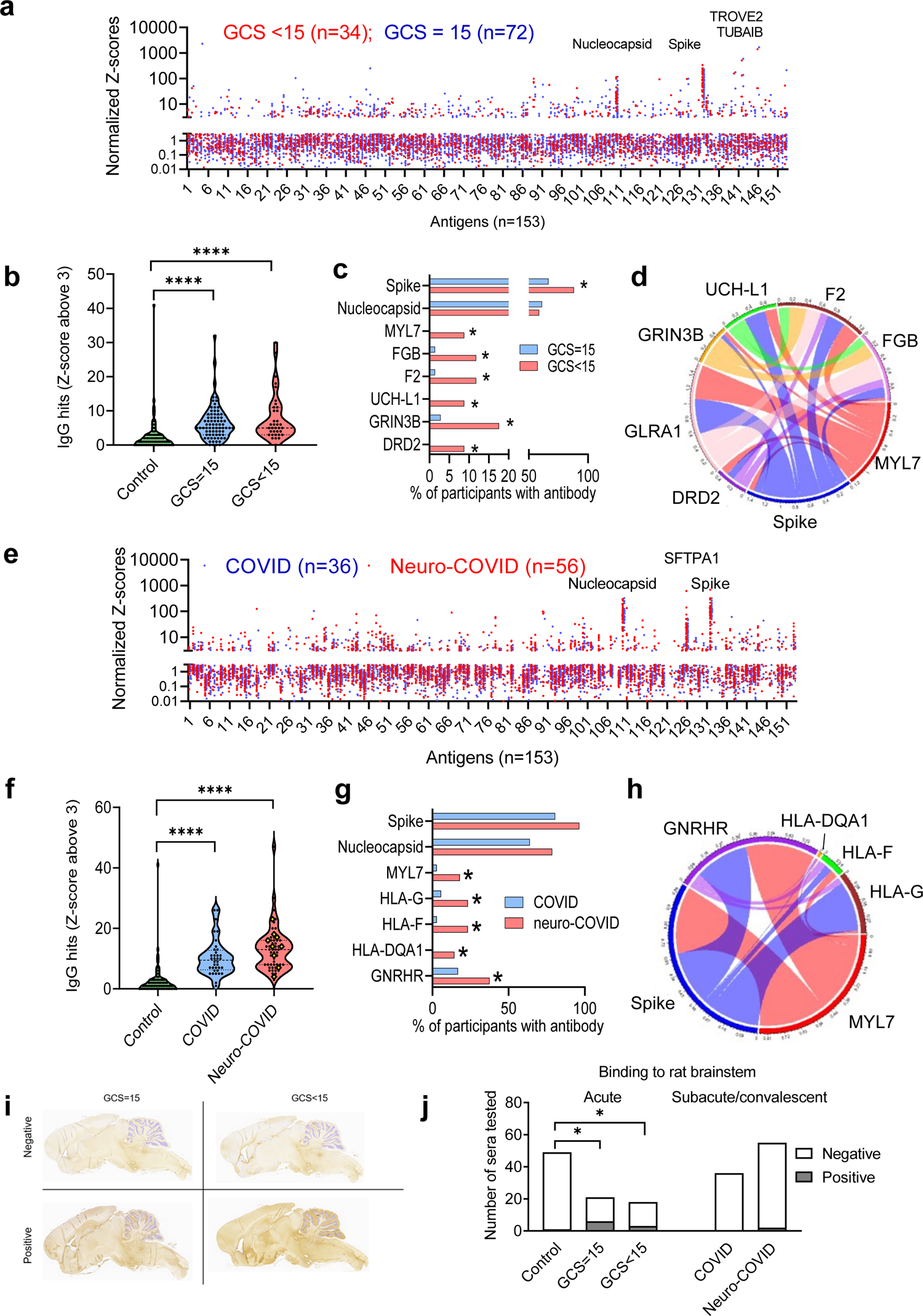
Acutely there is a polyclonal antibody response in patients with COVID-19 directed at SARS-CoV-2 spike protein and several CNS epitopes, including UCH-L1, and some correlate with NfL levels. **(a)** Acute samples were tested for IgG antibodies by protein microarray, normalized fluorescence Z-scores are shown. **(b)** COVID-19 patients showed considerably more binding ‘hits’ than controls (fluorescence with a Z-score of 3 or above compared to controls), although overall there was no difference in the acute samples between patients with normal or an abnormal GCS. **(c)** COVID-19 patients with abnormal GCS more frequently had several IgG antibodies than COVID-19 patients with a normal GCS, including those directed at SARS-CoV-2 spike protein and several CNS proteins (Fisher’s exact tests). **(d)** A Chord diagram shows the association of multiple antibodies with other antibodies, including anti-Spike antibodies **(e)** Sub-acute and convalescent serum was analysed for IgG antibodies by protein microarray. Normalized fluorescence Z-scores of IgG antibodies is shown. **(f)** A larger proportion of COVID-19+ve controls and cases had positive antibody ‘hits’ for IgG (defined by Z-score 3 and above compared to controls). **(g)** Of those antibodies against self-antigens identified, they were predominantly binding to human leucocyte antigen (HLA) epitopes (Fisher’s exact tests). At this timepoint there was no significant difference in the proportion of individuals with IgG against SARS-CoV-2 spike or nucleocapsid epitopes. **(i)** Representative images of rat brains incubated with participants’ sera and then stained with anti-human IgG to check for CNS-reactive antibodies **(j)|** Percentage of rat brain reactivity from serum of participants as detected by IHC— Fisher’s exact test with Benjamini and Hochberg correction. *Group comparisons are by Kruskal-Wallis test, pairwise comparisons by Mann-Whitney U test, and correlations are Pearson’s coefficients (* p<0.05, ** p<0.01, *** p<0.001, **** p<0.0001*).

Normalized fluorescence Z scores of serum autoantibodies in the subacute and convalescent samples were similar (Fig. 3e) to those in the acute samples (Fig. 3a), and the IgG ‘hits’ were more frequent than in controls, but were not different between COVID-19 patients with or without neurological complications (COVID and neuro-COVID, Fig. 3f, Extended Data Fig. 4a). However, specific autoantibody responses to MYL7, gonadotrophin releasing hormone receptor (GNRHR) and several HLA antigens had a frequency difference in the neuro-COVID participants (Fig. 3g, Extended Data Fig. 4b). As in the acute phase, autoantibody responses did not show significant associations with brain injury markers, but did tend to correlate with each other (Fig. 3h, Extended Data Fig. 4c)). Sera from acute COVID-19 participants with CNS antigen reactivity were incubated with sections of rat brain, neurons and antigen-expressing cells to explore binding to neuronal antigens. Binding to rat brain sections identified a small number of participants with strongly positive and characteristic immunohistochemical staining who might have CNS autoreactivity (Fig. 3i) and overall, sera from neuro-COVID participants showed more frequent binding to brainstem regions than control sera, but this did not relate to the GCS of the participants (Fig. 3j, Extended Data Fig. 5a).

**Figure 4:**
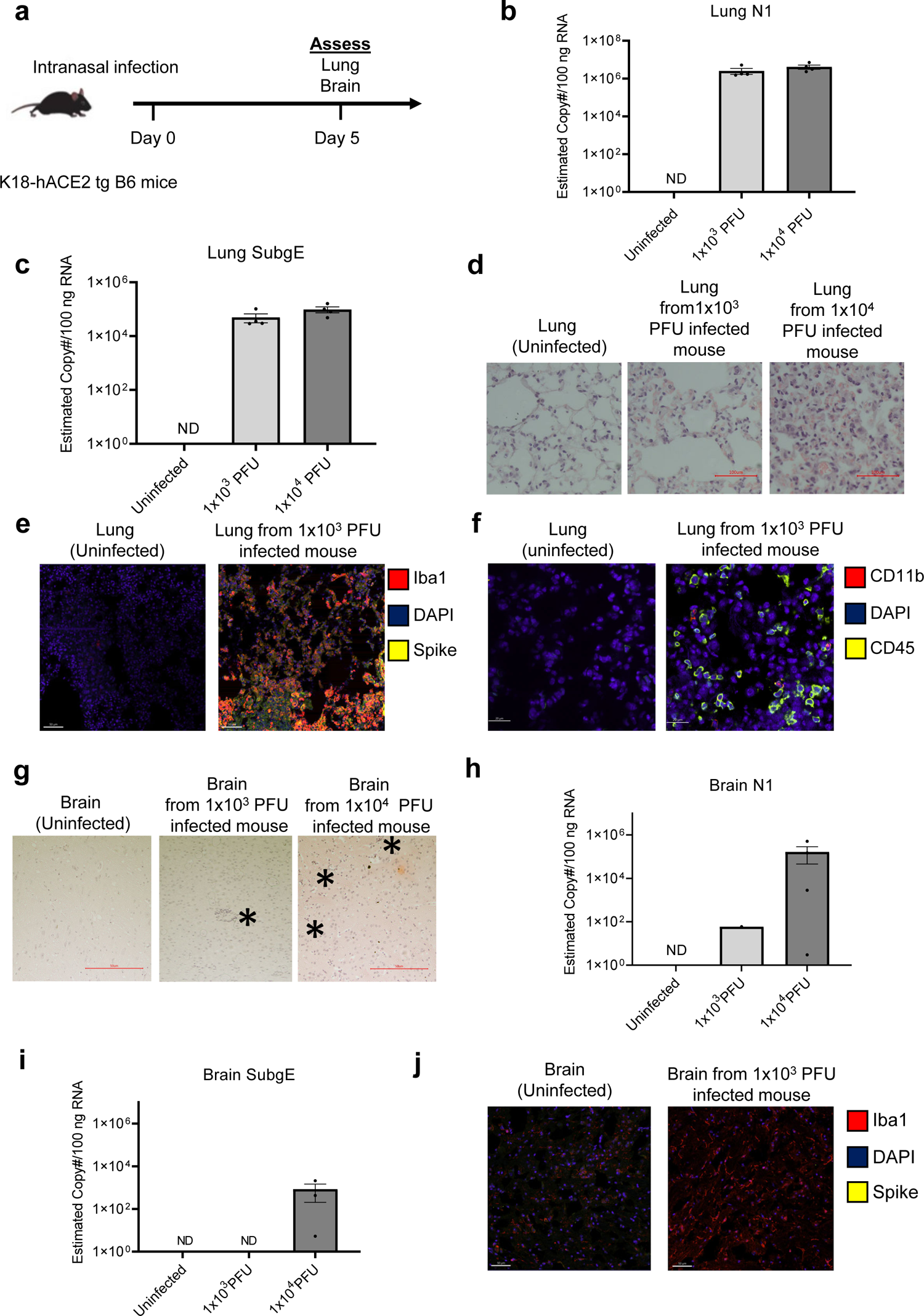
Low dose intranasal SARS-CoV-2 infection in human ACE-II transgenic C57BL/6 mice does not cause viral replication in the brain parenchyma, but does induce clustering of mononuclear cells, and microglial reactivity reflecting clinical disease. **(a)** Schema of K18 human-ACEII transgenic C57BL/6 mouse study randomised to no infection, ‘low inoculum’ infection at 1×10^3^ plaque-forming units (PFU) and ‘high inoculum’ infection at 1×10^4^ PFU, with endpoint at day 5 post-infection. **(b)** Real-time polymerase chain reaction (RT-QPCR) identifies SARS-CoV-2 N1 and **(c)** subgenomic E transcripts in lung homogenate confirming pulmonary infection and viral lytic replication at both inoculum of infection. **(d)** H&E staining of lung parenchyma demonstrates a dose-response relationship between control, low- and high-inoculum demonstrating an influx of mononuclear cells. **(e)** Confocal microscopy of low-inoculum infection in this model confirms the presence of both SARS-CoV-2 spike protein (yellow) and Iba1 positive cells of macrophage/monocyte lineage (red) shown with DAPI (blue). **(f)** Confocal microscopy of perfused lung in the low-inoculum SARS-CoV-2 infection model identifies the presence of CD45 positive leucocytes (yellow) and occasionally CD11b positive cells of macrophage/monocyte lineage (red), shown with DAPI (blue). **(g)** H&E staining of brain parenchyma from the frontal lobe demonstrates a dose response increase in mononuclear cells in both low and high-inoculum infection (asterisks). **(h)** RT-QPCR of brain homogenate demonstrates that minimal SARS-CoV-2 N1 is present in the perfused brain parenchyma and there is are no detectable SARS-CoV-2 subgenomic E transcripts **(i)** confirming the absence of lytic viral replication in the brain at low-dose infection. **(j)** Despite the absence of SARS-CoV-2 replication and spike protein in the brain in the low-dose infection model, there are consistent large areas of accumulation of Iba1+ microglia of perfused brain parenchyma.

**Figure 5:**
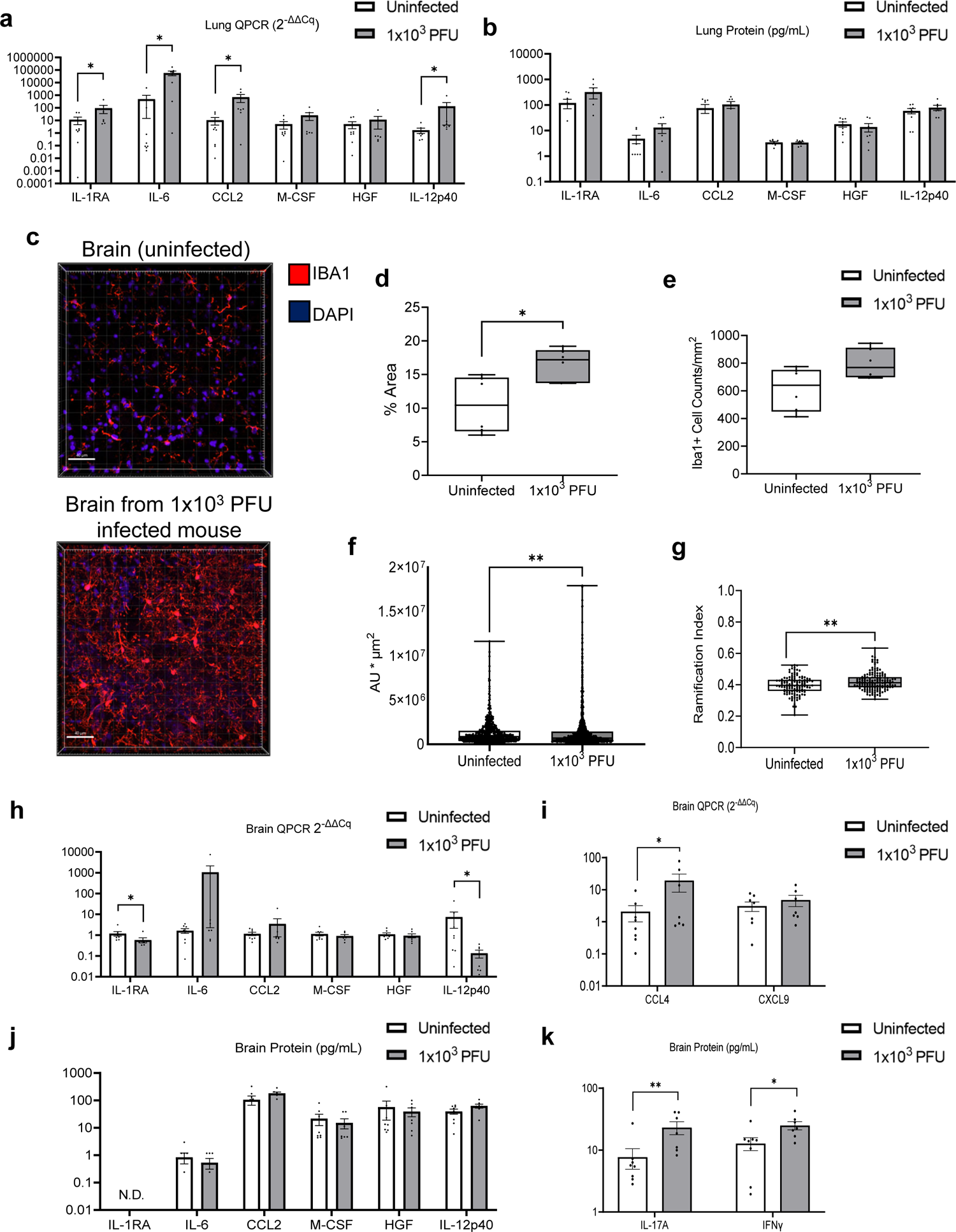
Low dose intranasal SARS-CoV-2 infection in mice results in increased pro-inflammatory cytokines in the lung and brain and causes increased microglia activation in the brain. **(a)** RT-QPCR of perfused lung parenchyma in the low-inoculum infection model (day 5 post 1×10^3^ PFU infection) confirms upregulation of several inflammatory mediators including CCL2, IL-6, and IL-12p40. **(b)** Luminex was used to check the same six immune mediators of interest in perfused lung in this model. **(c)** Confocal microscopy of brain parenchyma following low-inoculum infection reveals reactive microglia with increased expression of Iba1 (red) with nuclei stained by DAPI (purple). Reactive Iba1+ microglia were quantified by **(d)** % area that was Iba1-positive, **(e)** Iba1+ cell counts, **(f)** Iba1 fluorescence intensity, and **(g)** ramification index. **(h-k)** In this low-inoculum infection model several pro-inflammatory mediators were found to be increased in the perfused brains, including **(i)** CCL4 by RT-QPCR and **(k)** IL-17A and IFNγ by Luminex. *Pairwise comparisons by Mann-Whitney U test (* p<0.05, ** p<0.01, *** p<0.001, **** p<0.0001)*.

### Infection with low dose SARS-CoV-2 in a mouse model results in acute parainfectious brain pathology in the absence of active viral replication

Our results from the clinical samples suggested immune responses in COVID-19, some of which were associated with neurological injury (as defined by brain injury biomarker levels) or neurological dysfunction (defined by a reduced GCS or an acute COVID-19-related neurological complication). In order to further elucidate the mechanisms responsible for the acute neuroglial insult in COVID-19, we established a mouse model of para-infectious brain injury using intranasal infection with a low inoculum of SARS-CoV-2 (1×10^3^ PFU) and compared this to a high inoculum (1×10^4^ PFU); with assessment at day 5 post-infection (Fig. 4a). Both levels of infection caused pathology in the lung, with viral loads evidenced by qPCR of N1 in both low and high inocula of infection, but no weight loss (Fig. 4b, Extended Data Fig. 6a). Also, both resulted in evidence of active viral replication in the lung, as defined by qPCR of subgenomic E (Fig. 4c). Lung tissue from both high and low inoculum mice showed mononuclear cell infiltration which scaled with the severity of infection (Fig. 4d). When lung tissue from low-inoculum mice was examined by immunofluorescent staining and confocal microscopy, large amounts of viral protein were present and Iba1 expression was increased (Fig. 4e). These animals also showed increases in CD45 and CD11b staining in the lungs as opposed to uninfected animals (Fig. 4f, Extended Data Fig. 6b).

H&E staining of brain sections from infected mice showed clusters of mononuclear cells in the frontal cortex, which scaled in proportion to the dose of inoculum of SARS-CoV-2 (Fig. 4g; Extended Data Fig. 6c). SARS-CoV-2 N1 transcript was detected in the brain of mice which received high and low inoculum of SARS-CoV-2 in four out of five and six out of nine animals, respectively (Fig. 4h shows experiment one). However, whilst three of four mice which received the high inoculum showed active viral transcription in the brain, as evidenced by the detection of subgenomic E, this was only detectable in two of nine at the low inoculum (tissue from these two animals were not included in subsequent ex vivo experiments) (Fig. 4i). Confocal microscopy of ex vivo brain sections from mice following the low inoculum demonstrated Iba-1 staining but, in contrast to lung tissue, showed no evidence of staining for SARS-CoV-2 spike protein (Fig. 4j).

### Brains from low inoculum SARS-CoV-2 infected mice show increases in microglial and astrocyte activation despite the absence of active SARS-CoV-2 replication

In order to understand the mechanisms driving this para-infectious neuropathology, we assessed transcription and protein levels of inflammatory mediators in brains and lung from the seven low-inoculum infected mice that showed no evidence of local viral replication in the brain. Lung tissue revealed four out the six mediators of interest, as assessed by the clinical studies, to be increased with low-inoculum infection; IL-RA, IL-6, CCL2, and IL-12p40 (Fig. 5a,b). Interestingly, even though brains from these mice showed no detectable viral proteins (checked by spike staining) and no detectable viral transcription (checked using subgenomic E), these brains showed reactive microglia, with increased Iba1 expression and ramification indices (Fig. 5c-g). These brains showed no increases in numbers of CD45+, CD11b+, CD3+, and or NK1.1+ cells (Extended Data Fig. 6d,e,f), or apoptotic cells (Extended Data Fig. 6g). Interestingly, clusters of GFAP+ astrocytes were found in the regions of high Iba1 expression in brain, suggesting concomitant microgliosis and astrogliosis, as has been reported in human post-mortem samples (Extended Data Fig. 7a). Assessment of changes in inflammatory mediators by transcription, protein levels, and immunostaining yielded mixed results. The brains from low dose infected mice showed increased transcripts of CCL4 and decreased levels of IL-1RA and IL-12p40 (Fig. 5h,i) while both IFNγ and IL-17A were increased at the protein level in the low-inoculum infected mice (Fig. 5j,k). The brain/serum albumin ratio in low dose infected animals was higher than controls, but this did not achieve significance (Extended Data Fig. 7b). There were no significant differences in NfL, which may potentially be due to the early assessment timepoint (day 5 post infection) (both by ELISA and Simoa, Extended Data Fig. 7c).

## Discussion

We used several approaches to study neurological complications of COVID-19 infection. These included assessment of pathophysiology in participants with and without neurological complications, both in the acute and convalescent phases after COVID-19 infection; and examining systemic and neurological consequences in a para-infectious mouse model of SARS-CoV-2-associated encephalopathy. These complementary approaches allowed characterisation of pathophysiology and insights into pathogenesis. We demonstrated increased levels of brain injury biomarkers following COVID-19 infection, which showed specific patterns with disease phase (acute or convalescent), and varied with the presence or absence of neurological injury or dysfunction. In the acute phase, all four brain injury biomarkers (GFAP, NfL, Tau and UCH-L1) were elevated in participants when compared to controls, and specific markers of dendritic and axonal injury (Tau and NfL) were significantly higher in participants who showed a reduced level of consciousness (GCS≤14). In the subacute phase (<6 weeks post-infection), GFAP, NfL, and Tau were elevated in participants recovering from COVID-19, with no differences between those who had and had not sustained a neurological complication of disease. However, at late time points (>6 weeks) elevations NfL and GFAP were only seen in participants who had sustained a neurological complication of COVID-19 in the acute phase of their illness.

In the acute phase, when compared to controls, we also observed increases in a range of inflammatory mediators (IL-6, HGF, IL-12p40, IL-1RA, CCL2, and M-CSF) in the overall cohort of COVID-19 participants, with HGF and IL-12p40 showing robust differentiation between participants with and without alterations in consciousness. In contrast, participants at the late phase after COVID-19 infection showed no group level elevation of inflammatory mediators. However, late elevations in Tau correlated with levels of CCL2, CCL7, IL-1RA, IL-2Rα, M-CSF, SCF, IL-16, and IL-18, suggesting that these markers of the late innate host response were associated with persisting markers of dendritic/axonal injury markers. A network analysis showed that the repertoire of cytokine responses was different in participants both with acute reductions in GCS, or those recovering from a neurological complication of COVID-19 when compared to the GCS=15 group.

Participants with acute COVID-19 also developed IgG autoantibody responses to a significantly larger number of both neural and non-neural antigens, than seen in controls. These increased IgG responses persisted into the late phase. While the diversity of autoantibody response did not differ between participants with and without neurological dysfunction, autoantibody responses to specific antigens, including the neural antigens UCH-L1, GRIN3B, and DRD2, were more common in participants with abnormal GCS at presentation. In the late phase, participants who had and had not experienced a neurological complication of COVID-19 were distinguished by the presence of autoantibodies to HLA antigens in those with complications.

To better study these acute host response that drove these processes (and potentially resulted in neurological injury, dysfunction, and disease), and given that SARS-CoV-2 is rarely identified in the brain parenchyma in clinical samples, we developed a low-inoculum mouse model of COVID-19 which induced pulmonary infection in the absence of brain infection. In mice with pulmonary infection there was production of inflammatory mediators, including CCL2, IL-6, IL-12p40 and IL-1RA, in the lung. Despite the absence of viral replication in the brain parenchyma, there was some, albeit limited, production of inflammatory mediators in the brain, including CCL4, IFNγ and IL-17A. In addition to this, there was an increase in microglial Iba1 staining and colocalization of microglial and astrocyte clusters reflecting clinical disease driven by parainfectious processes. Taken together these findings suggest that a primarily pulmonary inflammatory process can drive parainfectious immune activation in the brain and the signature of an NK cell and/or T cell response, indicates a cascade of inflammation potentially amenable to treatment.

These data from clinical disease and a mouse model of low dose SARS-CoV-2 infection provide important insights regarding the pathophysiology and pathogenesis of neurological injury, dysfunction, and disease in COVID-19. The clinical characteristics of our participant cohorts, and the elevation in brain injury biomarkers, provide evidence of both acute and ongoing neurological injury. Further, the literature data on the rarity of direct CNS infection by the virus and the evidence from our para-infectious model of SARS-CoV-2 encephalopathy, suggest that the innate and adaptive host responses that we document should be explored as pathogenic mechanisms. The incidence of neurological cases has decreased since the first wave of the pandemic, possibly due to the use of immunosuppressants, such as dexamethasone, although this may also reflect vaccines attenuating disease and changes in the prevalence of different strains of SARS-CoV-2^23^.

The inflammatory mediators that we found to be elevated in the acute phase are broadly concordant with many other publications that have examined innate immune responses in COVID-19^18, 19^ but there are limited data addressing associations between such responses and the development of neurological complications. It is possible that some of the risk of developing such complications is simply related to the severity of systemic infection and the host response, and it would be surprising if this was not a strong contributor. However, our data suggest that acute neurological dysfunction in COVID-19 is also associated with a different repertoire of cytokine responses, with HGF and IL-12p40 showing the statistically most robust discrimination between participants with and without an abnormal GCS. HGF has important roles in brain development and synaptic biology^24^ and its elevation may represent a protective/reparative response in participants with neurological injury. IL-12p40 has a core role in orchestrating Th1 responses, and has been reported to be central in the development of central and peripheral neuroinflammation, with p40 monomer subunits perhaps acting as inhibitors of the process^25–27^. Interestingly, the cytokine network that was activated in the late convalescent phase was different, potentially indicating differential drivers of neurological injury throughout the disease course. Though group level comparisons with controls showed some commonalities in inflammatory mediator increase, most notably in IL-1RA, CCL2, and M-CSF there were many differences. The late Tau elevation that we demonstrated was significantly associated with elevations in these three mediators, but also CCL7, IL-2Rα, SCF, IL-16, and IL-18. These are all important pro-inflammatory mediators, and their association with Tau levels may reflect the persistence of a systemic inflammatory response that can enhance neuroinflammation^25, 27, 28^.

We found a polyclonal increase in antibody production following COVID-19 infection and only a few autoantibody frequencies were different when compared by GCS or COVID versus neuro-COVID cases. Of note, absolute levels of autoantibodies were low in comparison to anti-viral antibodies that developed over the course of the acute illness, with the exception of TROVE2, TUBA1B and SFTPA1. TROVE2, also known as Ro60 or SS-A is an RNA-binding protein and antibodies against it are present in autoimmune diseases including systemic lupus erythematosus and Sjögren’s syndrome, and its role in disease pathogenesis is an active area of research^29–31^. TUBA1B, a component of microtubules, has protein similarity to viral nucleocapsid and has been found to be recognized by antibodies from COVID-19 participant sera^32^. SFTPA1 is a lung surfactant protein and antibodies against it have been found to correlate with COVID-19 severity^12^. HLA alleles and even blood groups have been associated with COVID severity, but less is known about antibodies against them.^33, 34^. However, overall, we did not find a significant association of autoantibody levels with markers of brain injury, a finding that is against a broad causal role for these adaptive immune responses. Further analysis by screening the antibodies against brain antigens *ex vivo* revealed sporadic reactivity in both cases and controls with only the brainstem showing increased reactivity in COVID+ve participants but no difference between COVID and neuro-COVID.

Our studies have several limitations including: limited clinical information on the acute participants and lack of longitudinal blood samples; in addition, the low GCS could indicate sedation for intubation, rather than CNS disease, in the acute cohort. Although we did not have COVID-19 severity scores, we did know whether participants had required oxygen or not; when data were analysed within the cohorts just comparing participants who had required oxygen, 5 out of 6 cytokines remained significantly elevated in the abnormal GCS group. In the COVID-CNS study where we did have in-depth clinical information, again we were limited by not having prospective and longitudinal blood samples. Nevertheless, several cytokines showed significant positive correlations with the brain injury marker Tau, and interestingly, three of them were cytokines that were significantly associated with abnormal GCS in the acute cohort (IL-1RA, CCL2, and M-CSF) highlighting a network of co-upregulated biomarkers associated with neurological complications. This commonalities in innate immune response in patients who suffered neurological dysfunction/complications, both in the acute phase and at convalescence, is underlined by the results of network analysis. Pro-inflammatory cytokines are expected to be increased in the anti-viral response, but we found that they not only correlate with COVID-19 severity, but with GCS, as well.

Strengths of our study include the large cohort of participants studied with well-characterized neurological syndromes and a known range of timings since COVID-19 infection. We studied aspects of the innate and adaptive immune response as well as brain injury markers in order to discover useful biomarkers of neurological complications over time. Our mouse model is novel in using a low inoculum of SARS-CoV-2 virus for infection which does not involve lethal brain pathology, allowing us to study the immune activation in the brain in the absence of direct viral invasion. Our mouse model is congruent with the majority of human autopsy results which show limited virus in the brain, but nevertheless demonstrate inflammation and microglia reactivity^9,35^. It is worth noting that a hamster model which examined neuropathology at longer term follow-up (31 days post-SARS-CoV-2 infection) found that Iba1 expression remained elevated^36^. Studies of brain organoids have found that Iba1+ microglia engulf post-synaptic material contributing to synapse elimination^37^.

Several hypotheses for how SARS-CoV-2 causes neuropathology have been tested. A prospective study of hospitalised patients showing IL-6 and D-dimer as risk factors implicates the innate immune response and coagulation pathways^16^. The complement pathway and microthrombosis have been associated with brain endothelial damage from SARS-CoV-2 infection, and this phenotype persists months after COVID-19^35, 38^. Animal models have provided key insights into COVID-19 neuropathology that warrant discussion. There have been at least two reports of viral encephalitis and neuron degeneration and apoptosis observed in non-human primates^39, 40^. It is important to note that in these studies the virus was present at low amounts in the brain and predominantly in the vasculature as visualized by co-localization with Von Willebrand Factor^40^. Similar to the clinical scenario, there was no correlation of neuropathology with respiratory disease severity^33^. A recent mouse study is particularly relevant to our work and involved assessment of a mouse model that also lacked direct viral neural invasion by infecting mice that were intratracheally transfected with human ACE2. This study reported increased CXCL11 (eotaxin) in mouse serum and CSF which correlated with demyelination and this was recapitulated by giving CXCL11 intraperitoneally^41^. This was linked to clinical studies which showed elevated CXCL11 in patients with brain fog^41^. We did not observe elevated CXCL11 in our studies and this could be due to model and timepoint differences. Indeed, we did not observe any elevation of cytokines or brain injury markers in the sera of low dose infected mice (Extended Data Fig. 7c,d). However, it is very likely that these negative results are due to the protocols for heat-inactivation of experimental samples, which were part of safety protocols the CL3 lab^42^ at the point when these experiments were conducted. A combined analysis of hamster and clinical studies showed that COVID-19 infection led to IL-1β and IL-6 expression within the hippocampus and medulla oblongata and decreased neurogenesis in the hippocampal dentate gyrus which may relate to learning and memory deficits^43^. This was also borne out during *in vitro* studies that showed that serum from COVID+ve patients with delirium lead to decreased proliferation and increased apoptosis of a human hippocampal progenitor cell line mediated by elevated IL-6^44^.

In conclusion, we found that brain injury markers remain elevated in the sera of COVID-19 participants who have experienced acute neurological complications, with the highest levels observed in cerebrovascular and brain inflammation cases, with elevated levels persisting months after SARS-CoV-2 infection. If autoimmunity is involved in the neurological complications, it is not detectable in the sera at later timepoints by checking the pro-inflammatory cytokines elevated acutely, implicating local damage may be key. Our low inoculum SARS-CoV-2 mouse model highlights a way to study parainfectious effects on the brain and enables characterization of the brain tissue itself. The cytokine signature and microglia reactivity at day 5 post infection indicates an acute immune response including initial inflammation in the absence of active viral replication that could be amenable to targeted immunosuppression which can direct future studies.

## Methods

### Human participant studies/healthy controls and ethics information

The ISARIC WHO Clinical Characterization Protocol for Severe Emerging Infections in the UK (CCP-UK) was a prospective cohort study of hospitalised patients with COVID-19, which recruited across England, Wales, and Scotland (National Institute for Health Research Clinical Research Network Central Portfolio Management System ID: 14152) The protocol, revision history, case report form, patient information leaflets, consent forms and details of the Independent Data and Material Access Committee are available online^37^ For the ISARIC participants, we examined those participants with a normal vs. abnormal Glasgow coma score. In the COVID-CNS study (REC reference# 17/EE/0025), we could stratify by participants with neurological complications and specific conditions. Serum samples were collected at different timepoints from COVID-19 diagnosis. ISARIC samples were collected during the acute phase (1-11 days from admission). COVID-CNS samples were collected at outpatient follow up during the sub-acute (<6 weeks from admission) or convalescent (>6 weeks) phases. The samples were aliquoted, labelled with anonymised identifiers, and frozen immediately at −70°C.

### Human brain injury markers measurements

Brain injury markers were measured in sera using a Quanterix Simoa kit run on an automated HD-X Analyser according to the manufacturer’s protocol (Quanterix, Billerica, MA, USA, Neurology 4-Plex B Advantage Kit, cat#103345). We assessed neurofilament light chain (NfL), Ubiquitin C-Terminal Hydrolase L1 (UCH-L1), Tau, and glial fibrillary acidic protein (GFAP) in sera diluted 1:4 and used the manufacturer’s calibrators to calculate concentrations.

### Human serum cytokine measurements

Analytes were quantified using the BioRad human cytokine screening 48-plex kit (Cat# 12007283) following manufacturer’s instructions on a Bioplex 200 using Manager software 6.2. This involved incubation of 1:4 diluted sera with antibody-coated magnetic beads, automated magnetic plate washing, incubating the beads with secondary detection antibodies, and adding streptavidin-PE. Standard curves of known protein concentrations were used to quantify analytes. Samples that were under the limit of detection were valued at the lowest detectable value adjusted for 1:4 dilution factor.

### Median-centred normalization of human serum cytokine measurements

To minimise any potential impact of any possible variation in sample storage and transport, concentrations were median-centred and normalised for each participant, using established methodology^45–47^. The pg/mL of cytokines were log-transformed and the median per participant across all cytokines was calculated. The log-transformed median was subtracted from each log-transformed value to generate a normalized set.

### Protein microarray autoantibody profiling

Autoantibodies were measured from sera as previously described in Needham *et al*., 2022^12^. Briefly, a protein array of antigens (based on the HuProt™ (version 4.0) platform) was used to measure bound IgM and IgG from sera, using secondary antibodies with different fluorescent labels detected by a Tecan scanner. As developed in previous studies^12, 48^, antibody positivity was determined by measuring the median fluorescence intensity (MFI) of the four quadruplicate spots of each antigen. The MFI was then normalized to the MFI of all antigens for that participants’ sample by divided each value by the MFI. Z-scores were obtained from these normalized values based on the distribution derived for each antigen from the healthy control cohort. A positive autoantibody ‘hit’ was defined as an antigen where Z ≥ 3,.

### Immunohistochemistry

Immunohistochemistry was performed on sagittal sections of female Wistar rat brains. Brains were removed, fixed in 4% paraformaldehyde (PFA) at 4°C for 1 h, cryoprotected in 40% sucrose for 48 h, embedded in freezing medium and snap-frozen in isopentane chilled on dry ice. 10µm thick sections were cut and mounted on slides in a cryostat. A standard avidin-biotin peroxidase method was used, as reported previously^49, 50^, where sera were diluted 1:200 in 5% NGS and incubated at 4°C overnight, and secondary biotinylated goat anti-human IgG Fc was diluted (1:500) and incubated at room temperature for 1 h. Finally, slides were counter-stained using cresyl violet.

### Mouse studies of infection with SARS-CoV-2

An AWERB-approved protocol was followed for the two independent mouse studies. Male and female 2-4 month old heterozygote hACE2-transgenic C57BL/6 mice were intranasally infected with 1×10^3^ or 1×10^4^ plaque-forming units (PFU) of a human isolate of SARS-CoV2 (Liverpool Pango B). Mice were euthanized on day 5 post infection. Brains were perfused with 30mL PBS w/ 1mM EDTA, then one hemisphere fixed in formaldehyde-containing PLP buffer overnight at 4°C. Brains were then subjected to a sucrose gradient-- 10 and 20% sucrose for 1hr each and then 30% sucrose O/N at 4°C. Brains were then frozen in OCT by submerging moulds in a beaker of 2-methylbutane on dry ice. The other hemisphere was divided in two sagittal pieces and half preserved in 4%PFA for histology and half in trizol for RNA and protein extraction. Sera was collected and frozen and then heat-inactivated at 56°C for 30 mins prior to be moved from CL3 to CL2 lab. Lung tissue was preserved in 4%PFA for histology, in trizol for RNA and protein extraction, and in PLP for cryosectioning.

### qPCR of SARS-CoV-2 genes and mouse cytokines

Gene expression was measured from trizol isolated RNA (Invitrogen cat# 15596018, manufacturer’s protocol) using Promega’s GoTaq Probe 1-Step RT-qPCR system (cat#A6120, manufacturer’s protocol) on an Agilent AriaMx. Primers and FAM probes for SARS-CoV2, cytokines, and housekeeping genes were purchased from IDT (Extended Methods Table 1) with standard IDT qPCR primer/probe sets for the mouse cytokines.

The thermal cycle for N1 and the mouse cytokines was: 45°C for 15min 1x, 95°C for 2 min, then 45 cycles of 95°C for 3 secs followed by 55°C for 30 sec.

For subgenomic E: 45°C for 15 min 1x, 95°C for 2 min, then 45 cycles of 95°C for 15 secs followed by 58°C for 30 sec.

For 18S it was: 45°C for 15 min, 95°C for 2 min and 40 cycles of 95°C for 15s, 60°C for 1 min

**Extended Methods Table 1:**
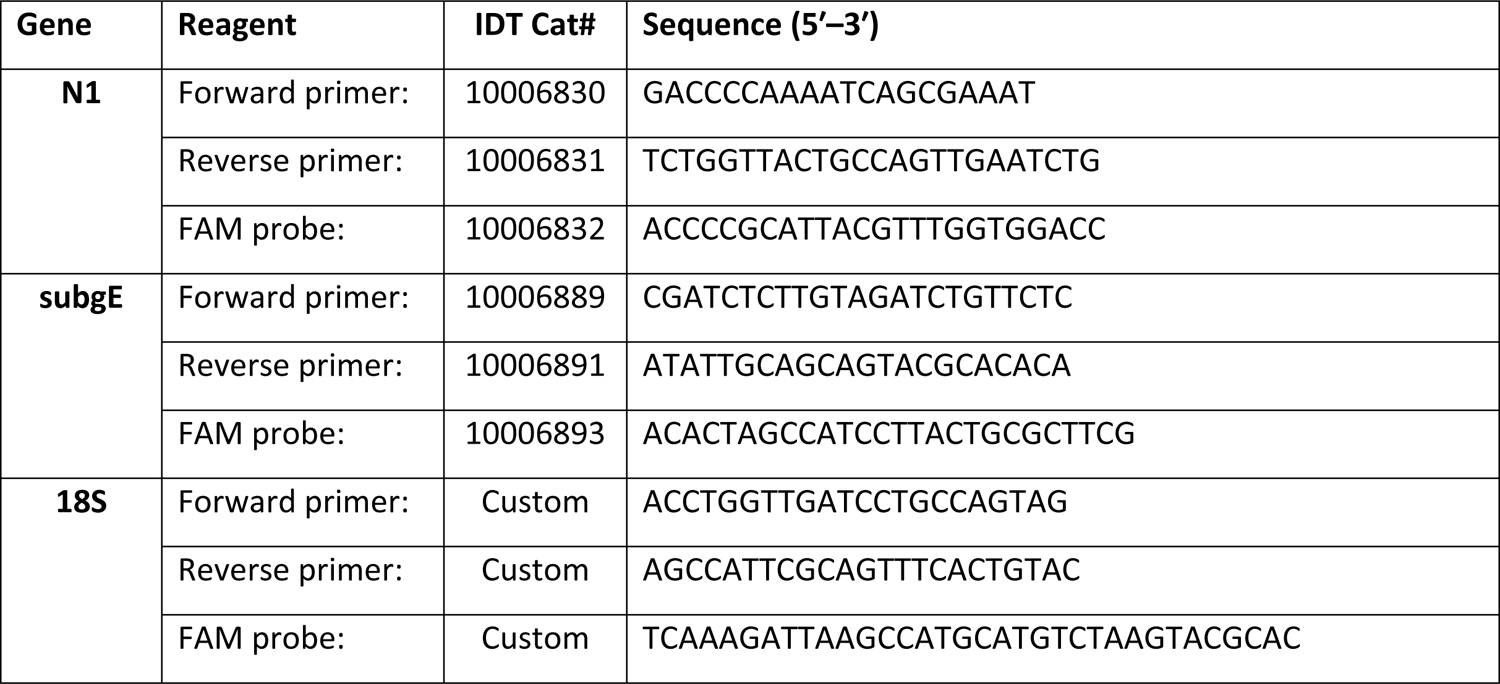
Primers and Probes for viral genes and normalization

**Extended Methods Table 2:** Primers and Probes for Mouse cytokines

| Gene | IDT Primetime Cat# |
| --- | --- |
| IL-1RN | Mm.PT.58.43781580 |
| IL-6 | Mm.PT.58.10005566 |
| IL-12p40 | Mm.PT.58.12409997 |
| M-CSF | Mm.PT.58.11661276 |
| CCL2 | Mm.PT.58.42151692 |
| HGF | Mm.PT.58.9088506 |
| CCL4 | Mm.PT.58.5219433 |
| IL-1RN (Exon 1-3) | Mm.PT.58.43781580 |
| IFNG | Mm.PT.58.41769240 |
| IL-17A | Mm.PT.58.6531092 |

For mouse cytokines, the primer/probe sets listed in Extended Methods Table 2 were used and the cycle was: 45°C for 15 min; 95°C for 2 min; 45 cycles of 95°C for 3 sec and 55°C for 30 sec.

### Mouse histology and confocal microscopy

The brain specificity of autoantibodies in participant sera were examined by assessing binding to mouse brain sections. Paraffin-embedded formalin-fixed tissue was sectioned to 4 um sections. Slides were baked at 60°C for 30 minutes and then stained with H&E in an autostainer. H&E slides were imaged on a Leica microsope (model details). For immunofluorescent staining and confocal microscopy, OCT-embedded frozen tissue was sectioned to 12 um sections. 100% acetone was used for antigen retrieval (10 mins at room temperature). After air-drying, then PBS washing, tissue sections were permeabilized with 0.1% Triton X-100/PBS (20 mins at room temperature). After rinsing with PBS, tissue sections were blocked with Dako block (5 minutes at room temperature). After another PBS wash, primary antibodies were added at dilutions listed in table for an overnight incubation at 4°C in a humidified chamber. Tissue sections were washed twice with PBS for 5 minutes each wash. Secondary antibody (as described in Extended Methods Table 3) was added for a 2 hr room temperature incubation, followed by two 5-minute PBS washes. DAPI-mounting medium was used for coverslipping. Imaging was performed with Andor Dragonfly spinning disk confocal microscope. Marker fluorescence and microglia counts, intensity, and ramification indices (method based on previous work^51^) were quantified with Fiji (confocal microscope set up in Extended Methods Table 4 and macros downloadable from the public <u>Github repository</u>). Ramification indices of 0-1 of objects with a threshold size of 19 mm^2^ were quantified from three Z-stack images/mouse and two mice/group.

**Extended Methods Table 3:** Antibodies for immunofluorescent stain and confocal imaging

| **Antibody target-fluorochrome** | **Company** | **Cat#** | **Host** | **Dilution Factor**<br>**(for 200 μL per section)** |
| --- | --- | --- | --- | --- |
| CD45-PE | Invitrogen | 12-0451-82 | Rat | 50 |
| CD11b-AF647 | BDBioscience | 557686 | Rat | 200 |
| NK1.1-AF488 | BioLegend | 108718 | Mouse | 100 |
| CD3-FITC | Abcam | ab34722 | Rat | 50 |
| GFAP | Invitrogen | 41-9892-82 | Mouse | 100 |
| NeuN | Merck | MAB377X | Mouse | 100 |
| CD68 unconjugated | Abcam | ab53444 | Rat | 200 |
| Anti-rat AF488 | Jackson Immunoresearch | 712-546-153 | Donkey | 500 |
| Iba1 unconjugated | Wako | 019-19741 | Rabbit | 500 |
| Anti-rabbit IgG-AF647 | Invitrogen | A-31573 | Donkey | 500 |
| Spike unconjugated | Invitrogen | 703958 | Human | 100 |
| Anti-human AF568 | Fisher | A-21090 | Goat | 500 |

**Extended Methods Table 4:** Confocal microscopy settings

| **Microscope component** | **Parameters** |
| --- | --- |
| Microscope | Leica DMI8 with Andor Dragonfly |
| Light source | 7-line integrated laser engine equipped with:<br>Solid state 405 smart diode laser at 100mW: set to 10%<br>Solid state 488 smart diode laser at 50mW: set to 35%<br>OBIS LS 561 smart OPSS laser at 50mW: set to 2.0%<br>OBIS LX Solid state 637 smart diode laser at 140mW: set to 7.0% |
| Excitation/emission optics | Dichroic mirror: Quad EM filter 405-488-561-640<br>Dual camera beam splitter: Dual camera dichroic 565nm long pass<br>Emission filters:<br>450/50nm bandpass filter<br>525/50nm bandpass filter<br>600/50nm bandpass filter<br>700/75nm bandpass filter<br>spinning disk with 40 μm pinholes |
| Objective lenses | Leica objectives:<br>11506358 HC PL APO 40×/1.30 OIL CS2 |
| Detector | Andor iXon Ultra 888 Ultra EMCCD Camera<br>1024 × 1024;<br>405: 500 ms exposure; 65 EM gain<br>488: 500 ms exposure; 65 EM gain<br>561: 30 ms exposure; 156 EM gain<br>637: 30 ms exposure; 156 EM gain<br>Averaging: 1; Binning: 1; camera magnification 1x<br>Nyquist Z sampling |

### Statistical Analyses

Prism software (version 9.4.1, GraphPad Software Inc.) was used for graph generation and statistical analysis. The Shapiro-Wilk normality test used to check the normality of the distribution. Data are expressed as mean ± S.E.M. Heatmaps, volcano plots and Chord diagrams were made using R studio (version 4.1.1 RStudio, PBC). The 2D cytokine network analyses were created using the qgraph package in R software and matrices differences were assessed by Steiger test^52^. Univariate analyses was conducted to test for differences between two groups. Differences between two normally distributed groups were tested using the paired or unpaired Student’s t test as appropriate. The difference between two non-normally distributed groups was tested using Mann-Whitney U test. Volcano plots used multiple Mann-Whitney U tests with a false discovery rate set to 5%, and heatmaps show Pearson’s correlations adjusted for a false discovery rate of 5%. Group comparisons were by Kruskal-Wallis test. Frequency differences of antibodies were measured by Fisher’s exact tests. P ≤ 0.05 was considered statistically significant.

## Data Availability

All data produced in the present study are available upon reasonable request to the authors.

https://github.com/Marien-kaefer/ramification_index

## Acknowledgements

BDM is supported by the UKRI/MRC (MR/V03605X/1), the MRC/UKRI (MR/V007181/1), MRC (MR/T028750/1) and Wellcome (ISSF201902/3). CD is supported by MRC (MC_PC_19044). We would like to thank the University of Liverpool GCP laboratory facility team for Luminex assistance and the Liverpool University Biobank team for all their help, especially Dr Victoria Shaw, Lara Lavelle-Langham, and Sue Holden. We would like to acknowledge the Liverpool Experimental Cancer Medicine Centre for providing infrastructure support for this research (Grant Reference: C18616/A25153). We acknowledge the Liverpool Centre for Cell Imaging (CCI) for provision of imaging equipment (Dragonfly confocal microscope) and excellent technical assistance (BBSRC grant number BB/R01390X/1). DKM and EN are supported by the NIHR Cambridge Biomedical Centre and by NIHR funding to the NIHR BioResource (RG94028 and RG85445), and by funding from Brain Research UK 201819-20.

---

**Extended data Table 1:**
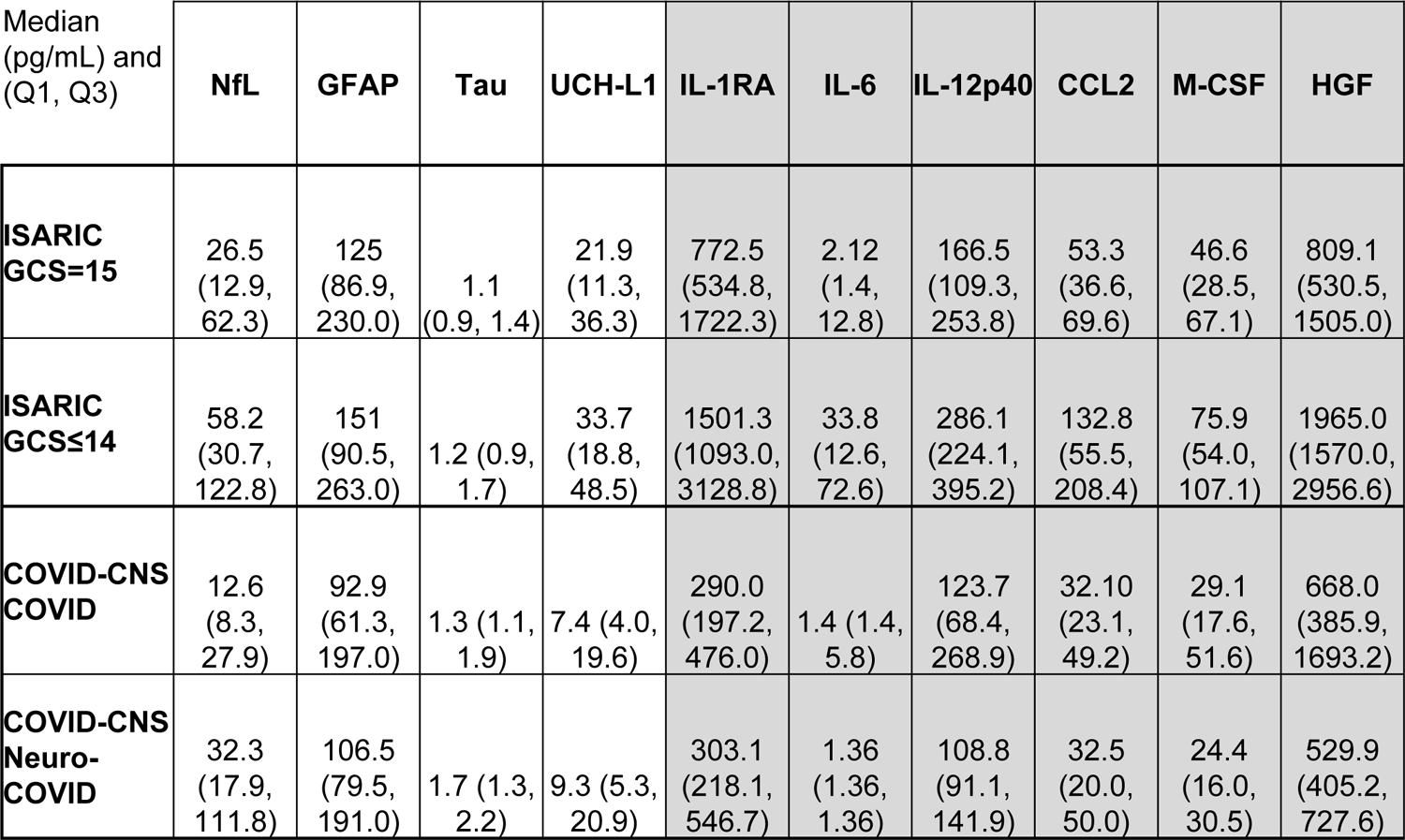
Median values of biomarkers

**Extended Data Table 2:**
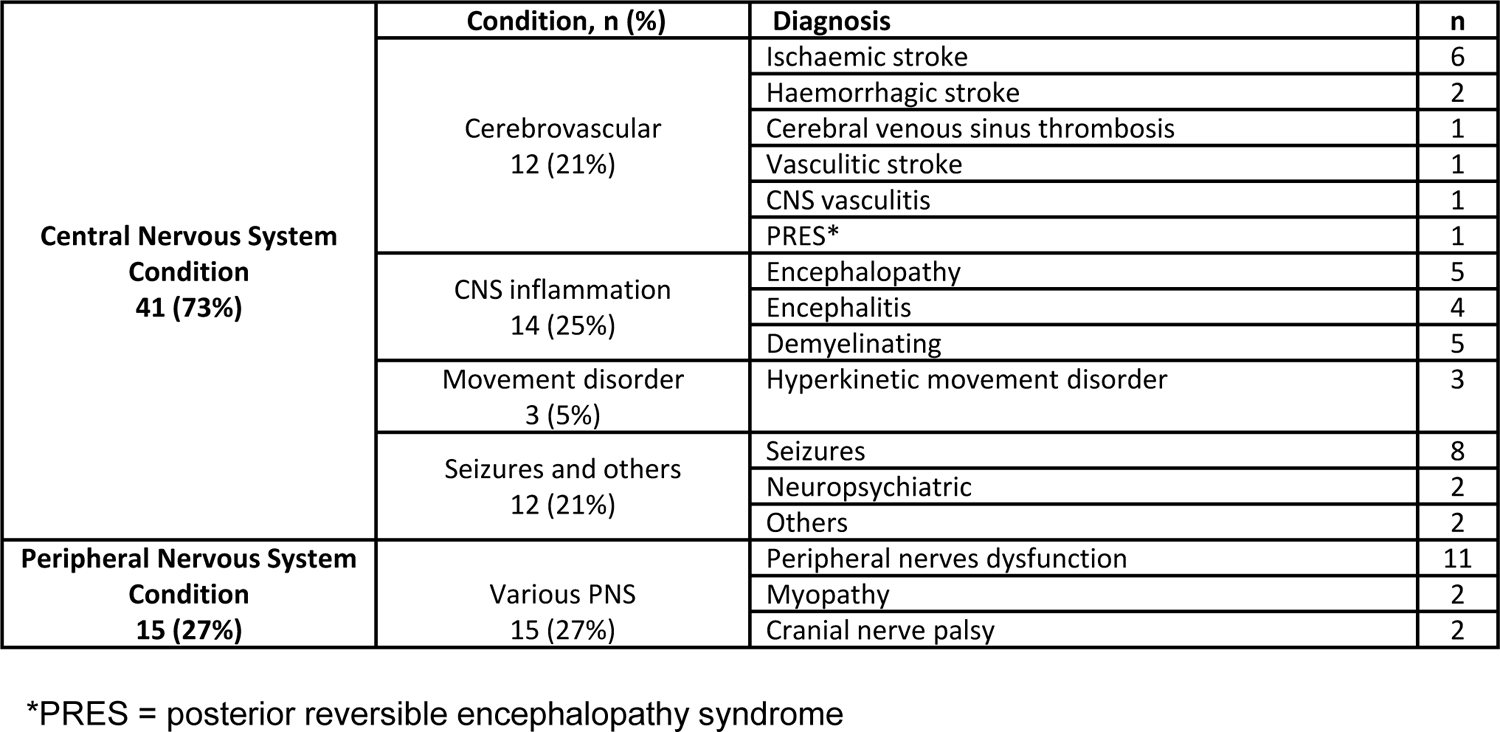
COVID-CNS Neuro-COVID cases

**Extended Data Table 3:**
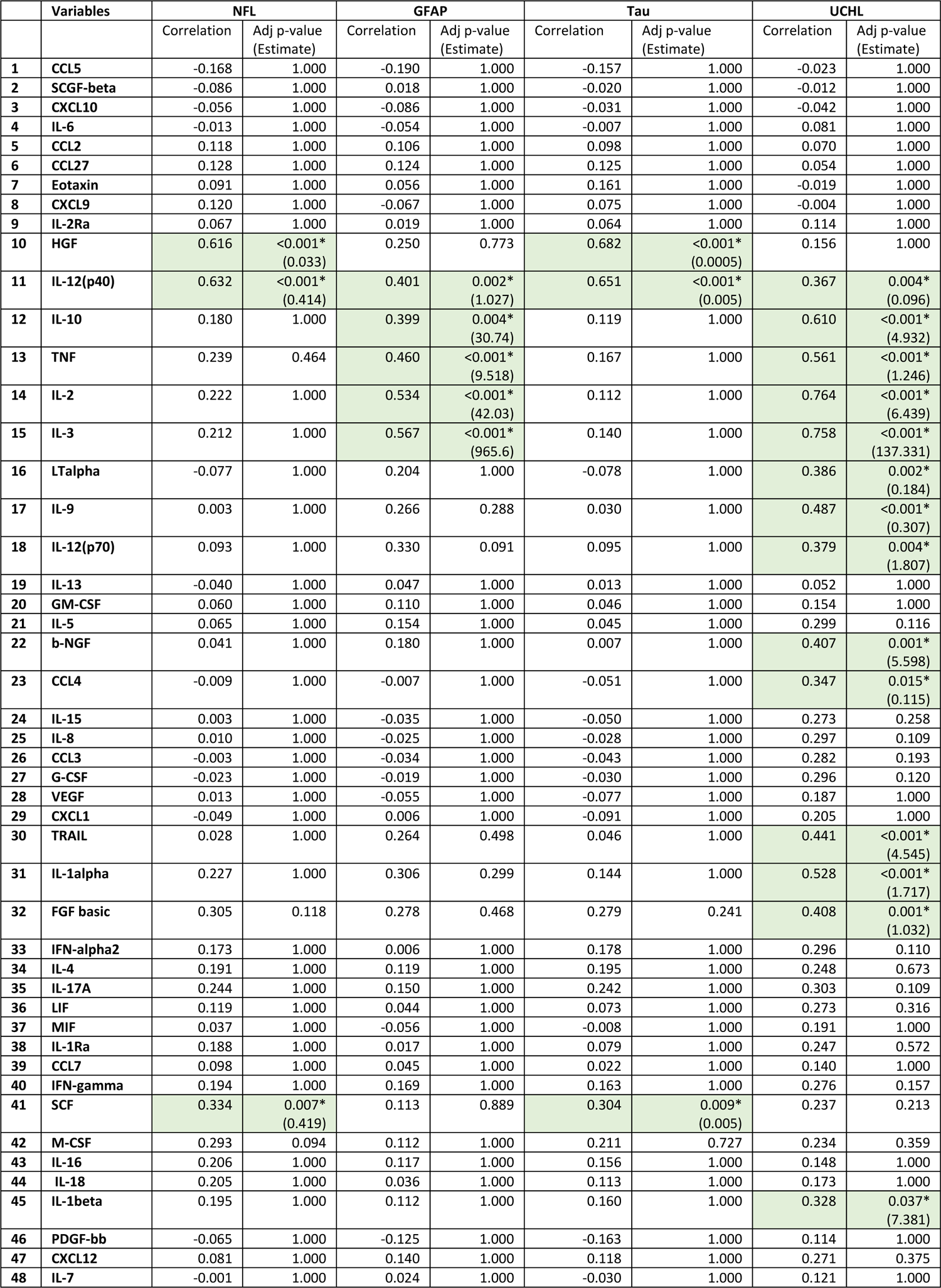
ISARIC cohort cytokines correlations With Brain injury markers

**Extended Data Table 4:**
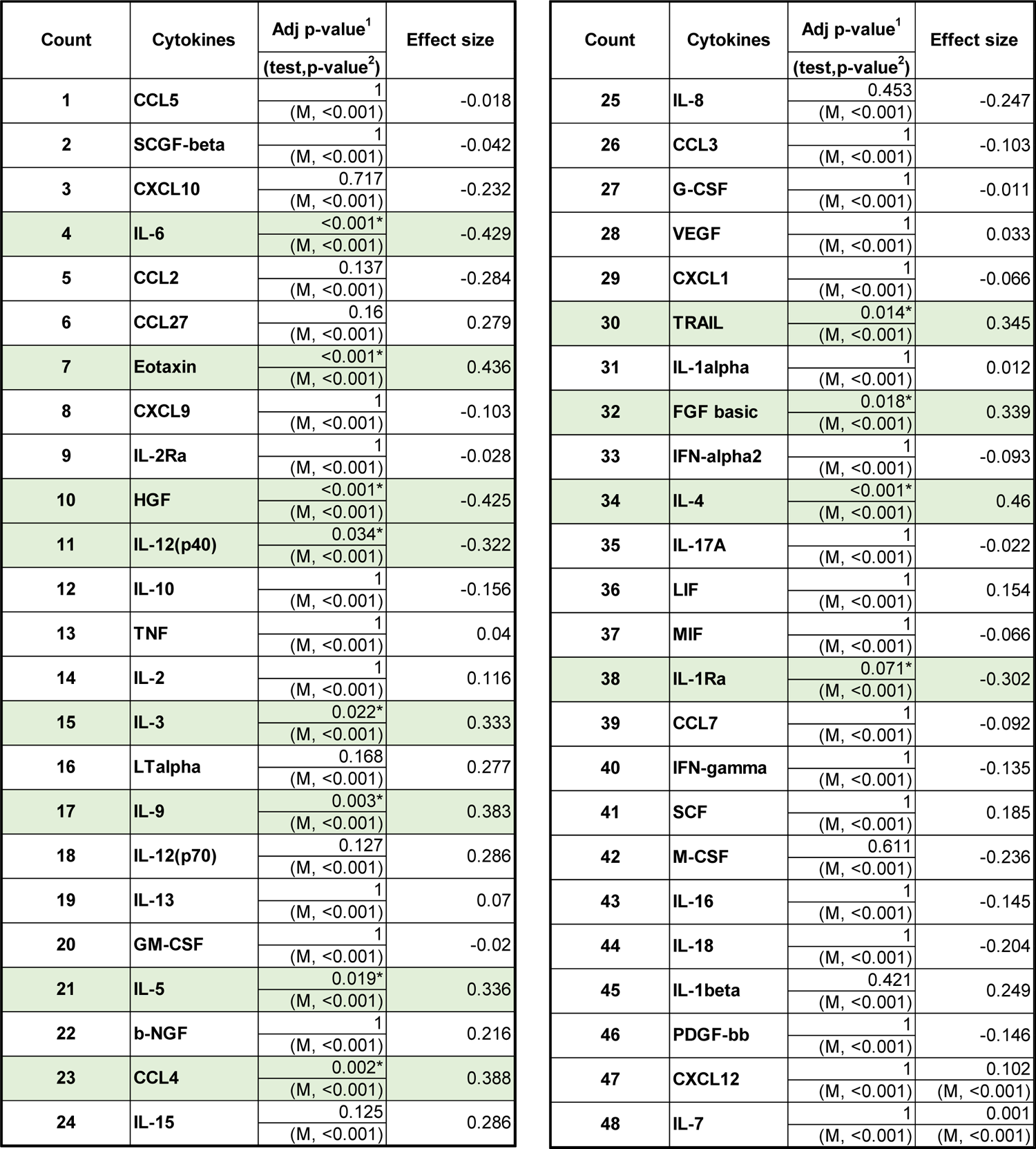
ISARIC Median-centred data Differences between GCS=15 and GCS≤14

| Count | Cytokines | Adj p-value<sup>1</sup> | Effect size |
| --- | --- | --- | --- |
|  |  | (test,p-value<sup>2</sup>) |  |
| 1 | CCL5 | 1<br>(M, <0.001) | -0.018 |
| 2 | SCGF-beta | 1<br>(M, <0.001) | -0.042 |
| 3 | CXCL10 | 0.717<br>(M, <0.001) | -0.232 |
| 4 | IL-6 | <0.001*<br>(M, <0.001) | -0.429 |
| 5 | CCL2 | 0.137<br>(M, <0.001) | -0.284 |
| 6 | CCL27 | 0.16<br>(M, <0.001) | 0.279 |
| 7 | Eotaxin | <0.001*<br>(M, <0.001) | 0.436 |
| 8 | CXCL9 | 1<br>(M, <0.001) | -0.103 |
| 9 | IL-2Ra | 1<br>(M, <0.001) | -0.028 |
| 10 | HGF | <0.001*<br>(M, <0.001) | -0.425 |
| 11 | IL-12(p40) | 0.034*<br>(M, <0.001) | -0.322 |
| 12 | IL-10 | 1<br>(M, <0.001) | -0.156 |
| 13 | TNF | 1<br>(M, <0.001) | 0.04 |
| 14 | IL-2 | 1<br>(M, <0.001) | 0.116 |
| 15 | IL-3 | 0.022*<br>(M, <0.001) | 0.333 |
| 16 | LTalpha | 0.168<br>(M, <0.001) | 0.277 |
| 17 | IL-9 | 0.003*<br>(M, <0.001) | 0.383 |
| 18 | IL-12(p70) | 0.127<br>(M, <0.001) | 0.286 |
| 19 | IL-13 | 1<br>(M, <0.001) | 0.07 |
| 20 | GM-CSF | 1<br>(M, <0.001) | -0.02 |
| 21 | IL-5 | 0.019*<br>(M, <0.001) | 0.336 |
| 22 | b-NGF | 1<br>(M, <0.001) | 0.216 |
| 23 | CCL4 | 0.002*<br>(M, <0.001) | 0.388 |
| 24 | IL-15 | 0.125<br>(M, <0.001) | 0.286 |
|  |  | (test,p-value<sup>2</sup>) |  |
| 25 | IL-8 | 0.453<br>(M, <0.001) | -0.247 |
| 26 | CCL3 | 1<br>(M, <0.001) | -0.103 |
| 27 | G-CSF | 1<br>(M, <0.001) | -0.011 |
| 28 | VEGF | 1<br>(M, <0.001) | 0.033 |
| 29 | CXCL1 | 1<br>(M, <0.001) | -0.066 |
| 30 | TRAIL | 0.014*<br>(M, <0.001) | 0.345 |
| 31 | IL-1alpha | 1<br>(M, <0.001) | 0.012 |
| 32 | FGF basic | 0.018*<br>(M, <0.001) | 0.339 |
| 33 | IFN-alpha2 | 1<br>(M, <0.001) | -0.093 |
| 34 | IL-4 | <0.001*<br>(M, <0.001) | 0.46 |
| 35 | IL-17A | 1<br>(M, <0.001) | -0.022 |
| 36 | LIF | 1<br>(M, <0.001) | 0.154 |
| 37 | MIF | 1<br>(M, <0.001) | -0.066 |
| 38 | IL-1Ra | 0.071*<br>(M, <0.001) | -0.302 |
| 39 | CCL7 | 1<br>(M, <0.001) | -0.092 |
| 40 | IFN-gamma | 1<br>(M, <0.001) | -0.135 |
| 41 | SCF | 1<br>(M, <0.001) | 0.185 |
| 42 | M-CSF | 0.611<br>(M, <0.001) | -0.236 |
| 43 | IL-16 | 1<br>(M, <0.001) | -0.145 |
| 44 | IL-18 | 1<br>(M, <0.001) | -0.204 |
| 45 | IL-1beta | 0.421<br>(M, <0.001) | 0.249 |
| 46 | PDGF-bb | 1<br>(M, <0.001) | -0.146 |
| 47 | CXCL12 | 1<br>(M, <0.001) | 0.102<br>(M, <0.001) |
| 48 | IL-7 | 1<br>(M, <0.001) | 0.001<br>(M, <0.001) |

**Extended Data Table 5:**
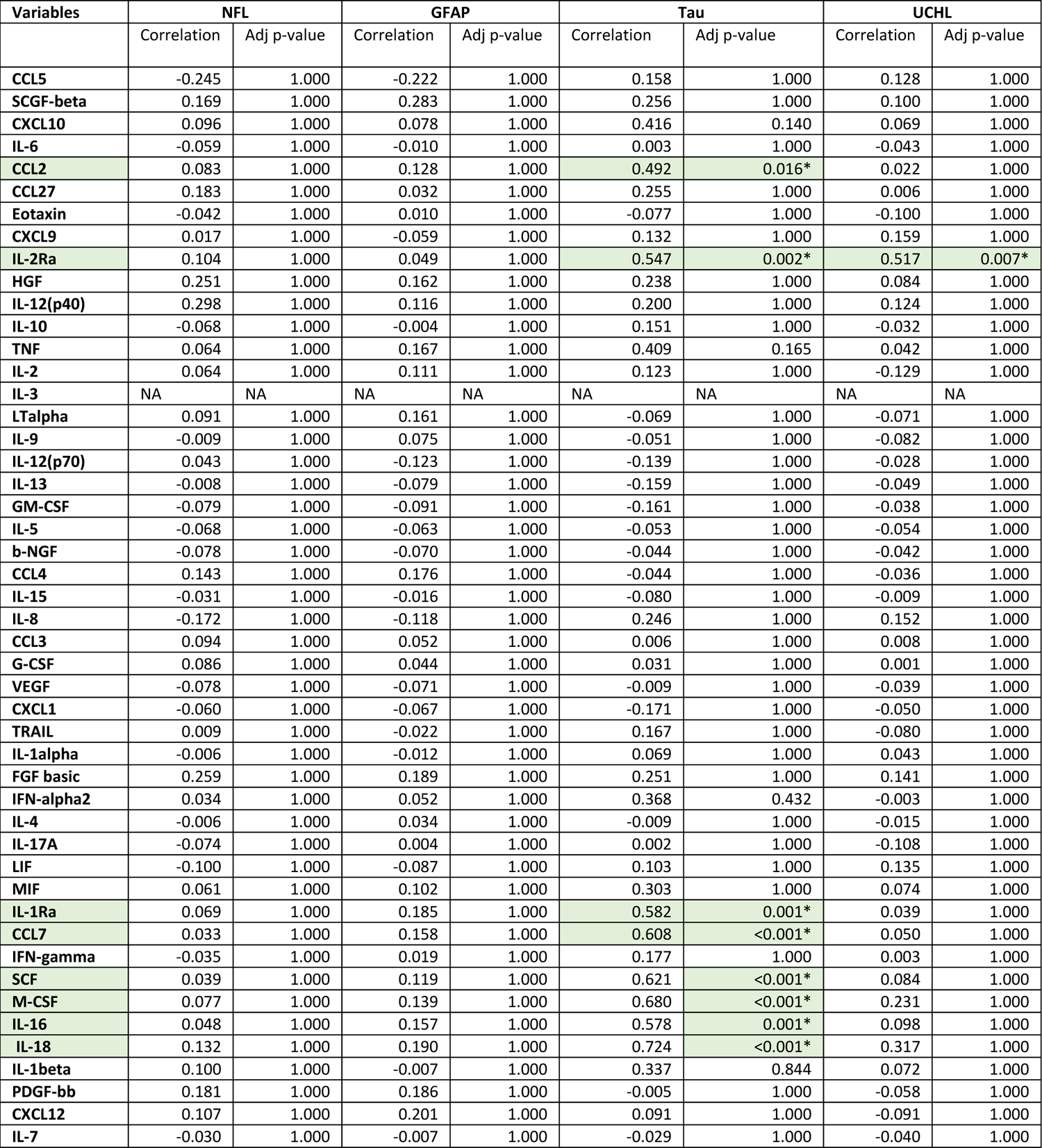
Correlations of cytokines and brain injury markers in the COVID-CNS Neuro-COVID cohort with regression models

**Extended Data Table 6:**
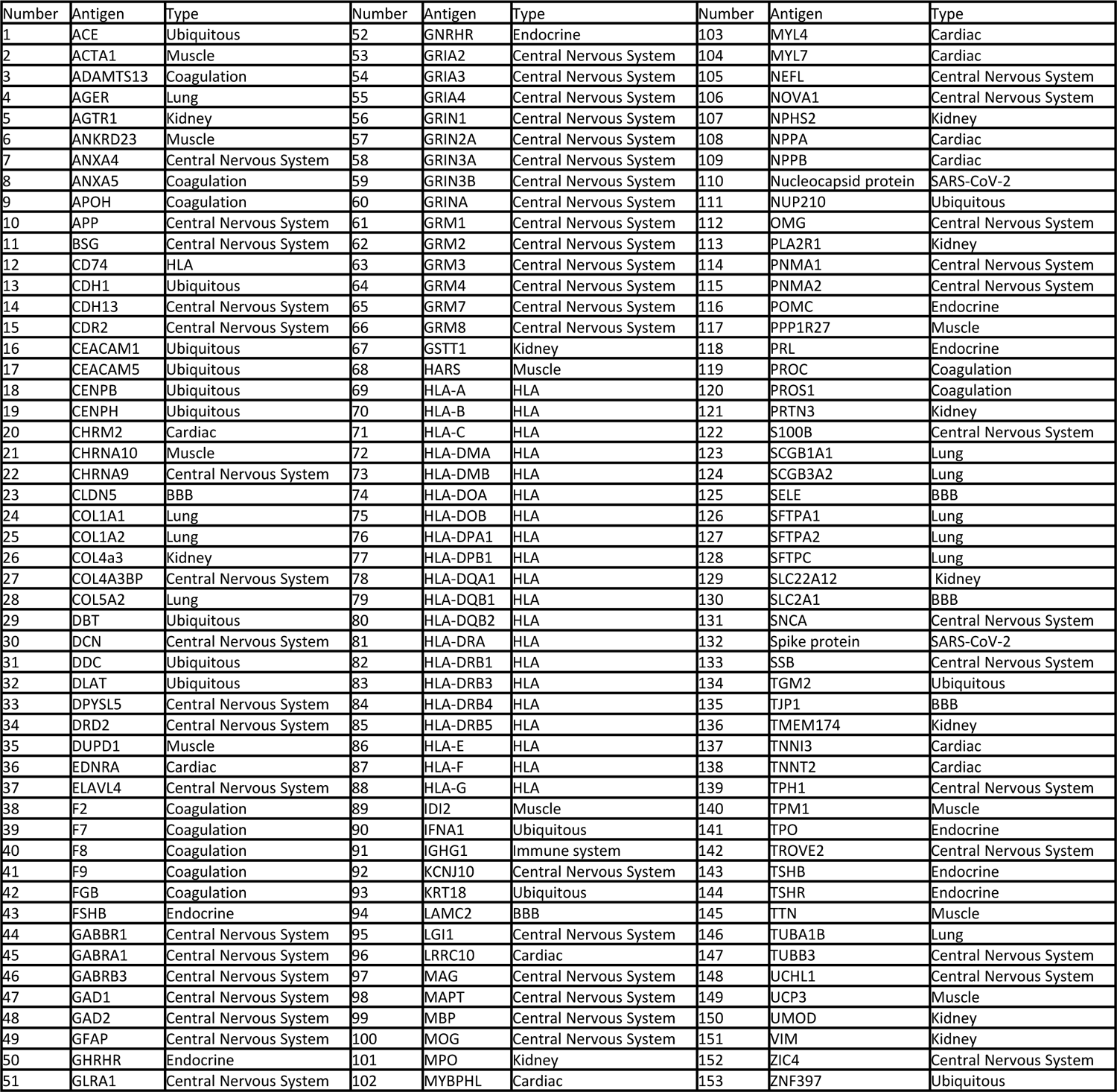
List of antigens on protein array

## References

1. Mao, L. et al. Neurologic Manifestations of Hospitalized Patients With Coronavirus Disease 2019 in Wuhan, China. JAMA Neurol. 77, 683–690 (2020).

2. Xu, E., Xie, Y. & Al-Aly, Z. Long-term neurologic outcomes of COVID-19. Nat. Med. 1–10 (2022)doi:10.1038/s41591-022-02001-z.

3. Varatharaj, A. et al. Neurological and neuropsychiatric complications of COVID-19 in 153 patients: a UK-wide surveillance study. Lancet Psychiatry 7, 875–882 (2020).

4. Ross Russell, A. L., et al. Spectrum, risk factors and outcomes of neurological and psychiatric complications of COVID-19: a UK-wide cross-sectional surveillance study. Brain Commun. 3, fcab168 (2021).

5. Paterson, R. W. et al. The emerging spectrum of COVID-19 neurology: clinical, radiological and laboratory findings. Brain 143, 3104–3120 (2020).

6. Song, E. et al. Neuroinvasion of SARS-CoV-2 in human and mouse brain. J. Exp. Med. 218, e20202135 (2021).

7. Morphological, cellular, and molecular basis of brain infection in COVID-19 patients. https://www.pnas.org/doi/10.1073/pnas.2200960119 doi:10.1073/pnas.2200960119.

8. Meinhardt, J. et al. Olfactory transmucosal SARS-CoV-2 invasion as a port of central nervous system entry in individuals with COVID-19. Nat. Neurosci. 24, 168–175 (2021).

9. Thakur, K. T., et al. COVID-19 neuropathology at Columbia University Irving Medical Center/New York Presbyterian Hospital. Brain 144, 2696–2708 (2021).

10. Khan, M. et al. Visualizing in deceased COVID-19 patients how SARS-CoV-2 attacks the respiratory and olfactory mucosae but spares the olfactory bulb. Cell 184, 5932–5949.e15 (2021).

11. Domingues, R. B., Leite, F. B. V. de M. & Senne, C. Cerebrospinal fluid analysis in patients with COVID-19-associated central nervous system manifestations: a systematic review. Arq. Neuropsiquiatr. S0004-282X2022005004205 (2022) doi:10.1590/0004-282X-ANP-2021-0117.

12. Needham, E. J. et al. Brain injury in COVID-19 is associated with dysregulated innate and adaptive immune responses. Brain awac321 (2022) doi:10.1093/brain/awac321.

13. Dunai, C., Collie, C. & Michael, B. D. Immune-Mediated Mechanisms of COVID-19 Neuropathology. Front. Neurol. 13, (2022).

14. Maiese, A. et al. SARS-CoV-2 and the brain: A review of the current knowledge on neuropathology in COVID-19. Brain Pathol. 31, e13013 (2021).

15. Fara, M. G. et al. Macrothrombosis and stroke in patients with mild Covid-19 infection. J. Thromb. Haemost. 18, 2031–2033 (2020).

16. Frontera, J. A. et al. A Prospective Study of Neurologic Disorders in Hospitalized Patients With COVID-19 in New York City. Neurology 96, e575–e586 (2021).

17. Hampshire, A. et al. Multivariate profile and acute-phase correlates of cognitive deficits in a COVID-19 hospitalised cohort. eClinicalMedicine 47, 101417 (2022).

18. Lucas, C. et al. Longitudinal analyses reveal immunological misfiring in severe COVID-19 Nature 584, 463–469 (2020).

19. Thwaites, R. S., et al. Inflammatory profiles across the spectrum of disease reveal a distinct role for GM-CSF in severe COVID-19. Sci. Immunol. 6, eabg9873 (2021).

20. Perreau, M. et al. The cytokines HGF and CXCL13 predict the severity and the mortality in COVID-19 patients. Nat. Commun. 12, 4888 (2021).

21. Li, L. et al. Interleukin-8 as a Biomarker for Disease Prognosis of Coronavirus Disease-2019 Patients. Front. Immunol. 11, (2021).

22. Bertin, D. et al. Anticardiolipin IgG Autoantibody Level Is an Independent Risk Factor for COVID-19 Severity. Arthritis Rheumatol. Hoboken Nj 72, 1953–1955 (2020).

23. Grundmann, A. et al. Impact of Dexamethasone and Remdesivir on Neurological Complications during COVID-19. SSRN Scholarly Paper at https://doi.org/10.2139/ssrn.4065552 (2022).

24. Desole, C. et al. HGF and MET: From Brain Development to Neurological Disorders. Front. Cell Dev. Biol. 9, (2021).

25. Bao, L. et al. The Critical Role of IL-12p40 in Initiating, Enhancing, and Perpetuating Pathogenic Events in Murine Experimental Autoimmune Neuritis. Brain Pathol. 12, 420–429 (2002).

26. Mondal, S. et al. IL-12 p40 monomer is different from other IL-12 family members to selectively inhibit IL-12Rβ1 internalization and suppress EAE. Proc. Natl. Acad. Sci. 117, 21557–21567 (2020).

27. Kroenke, M. A., Carlson, T. J., Andjelkovic, A. V. & Segal, B. M. IL-12- and IL-23-modulated T cells induce distinct types of EAE based on histology, CNS chemokine profile, and response to cytokine inhibition. J. Exp. Med. 205, 1535–1541 (2008).

28. Yu, S. et al. Neutralizing Antibodies to IL-18 Ameliorate Experimental Autoimmune Neuritis by Counter-Regulation of Autoreactive Th1 Responses to Peripheral Myelin Antigen. J. Neuropathol. Exp. Neurol. 61, 614–622 (2002).

29. Boccitto, M. & Wolin, S. L. Ro60 and Y RNAs: structure, functions, and roles in autoimmunity. Crit. Rev. Biochem. Mol. Biol. 54, 133–152 (2019).

30. Zampeli, E., Mavrommati, M., Moutsopoulos, H. M. & Skopouli, F. N. Anti-Ro52 and/or anti-Ro60 immune reactivity: autoantibody and disease associations. Clin. Exp. Rheumatol. 8 (2020).

31. Hung, T. et al. The Ro60 autoantigen binds endogenous retroelements and regulates inflammatory gene expression. Science 350, 455–459 (2015).

32. Matyushkina, D. et al. Autoimmune Effect of Antibodies against the SARS-CoV-2 Nucleoprotein. Viruses 14, 1141 (2022).

33. Palmos, A. B. et al. Proteome-wide Mendelian randomization identifies causal links between blood proteins and severe COVID-19. PLOS Genet. 18, e1010042 (2022).

34. Tavasolian, F. et al. HLA, Immune Response, and Susceptibility to COVID-19. Front. Immunol. 11, (2021).

35. Lee, M. H. et al. Neurovascular injury with complement activation and inflammation in COVID-19. Brain J. Neurol. 145, 2555–2568 (2022).

36. Frere, J. J. et al. SARS-CoV-2 infection in hamsters and humans results in lasting and unique systemic perturbations after recovery. Sci. Transl. Med. 14, eabq3059 (2022).

37. Samudyata et al. SARS-CoV-2 promotes microglial synapse elimination in human brain organoids. Mol. Psychiatry 27, 3939–3950 (2022).

38. Pretorius, E. et al. Persistent clotting protein pathology in Long COVID/Post-Acute Sequelae of COVID-19 (PASC) is accompanied by increased levels of antiplasmin. Cardiovasc. Diabetol. 20, 172 (2021).

39. Choudhary, S. et al. Modeling SARS-CoV-2: Comparative Pathology in Rhesus Macaque and Golden Syrian Hamster Models. Toxicol. Pathol. 01926233211072767 (2022) doi:10.1177/01926233211072767.

40. Rutkai, I. et al. Neuropathology and virus in brain of SARS-CoV-2 infected non-human primates. Nat. Commun. 13, 1745 (2022).

41. Fernández-Castañeda, A. et al. Mild respiratory SARS-CoV-2 infection can cause multi-lineage cellular dysregulation and myelin loss in the brain. 2022.01.07.475453 Preprint at https://www.biorxiv.org/content/10.1101/2022.01.07.475453v1 (2022).

42. Xu, E., Li, T., Chen, Q., Wang, Z. & Xu, Y. Study on the Effect and Application Value of Heat-Inactivated Serum on the Detection of Thyroid Function, Tumor Markers, and Cytokines During the SARS-CoV-2 Pandemic. Front. Med. 8, (2021).

43. Soung, A. L. et al. COVID-19 induces CNS cytokine expression and loss of hippocampal neurogenesis. Brain awac270 (2022) doi:10.1093/brain/awac270.

44. Borsini, A. et al. Neurogenesis is disrupted in human hippocampal progenitor cells upon exposure to serum samples from hospitalized COVID-19 patients with neurological symptoms. Mol. Psychiatry 1–13 (2022) doi:10.1038/s41380-022-01741-1.

45. Michael, B. D. et al. Characteristic Cytokine and Chemokine Profiles in Encephalitis of Infectious, Immune-Mediated, and Unknown Aetiology. PLoS ONE 11, e0146288 (2016).

46. Griffiths, M. J. et al. In Enterovirus 71 Encephalitis With Cardio-Respiratory Compromise, Elevated Interleukin 1β, Interleukin 1 Receptor Antagonist, and Granulocyte Colony-Stimulating Factor Levels Are Markers of Poor Prognosis. J. Infect. Dis. 206, 881– 892 (2012).

47. Michael, B. D. et al. Post-acute serum eosinophil and neutrophil-associated cytokine/chemokine profile can distinguish between patients with neuromyelitis optica and multiple sclerosis; and identifies potential pathophysiological mechanisms – A pilot study. Cytokine 64, 90–96 (2013).

48. Needham, E. J. et al. Complex Autoantibody Responses Occur following Moderate to Severe Traumatic Brain Injury. J. Immunol. 207, 90–100 (2021).

49. Ances, B. M. et al. Treatment-responsive limbic encephalitis identified by neuropil antibodies: MRI and PET correlates. Brain 128, 1764–1777 (2005).

50. Lai, M. et al. Investigation of LGI1 as the antigen in limbic encephalitis previously attributed to potassium channels: a case series. Lancet Neurol. 9, 776–785 (2010).

51. Wittekindt, M. et al. Different Methods for Evaluating Microglial Activation Using Anti-Ionized Calcium-Binding Adaptor Protein-1 Immunohistochemistry in the Cuprizone Model. Cells 11, 1723 (2022).

52. Masi, A. et al. Cytokine levels and associations with symptom severity in male and female children with autism spectrum disorder. Mol. Autism 8, 63 (2017).

