## Extended Data for "Para-infectious brain injury in COVID-19 persists at follow-up despite attenuated cytokine and autoantibody responses"

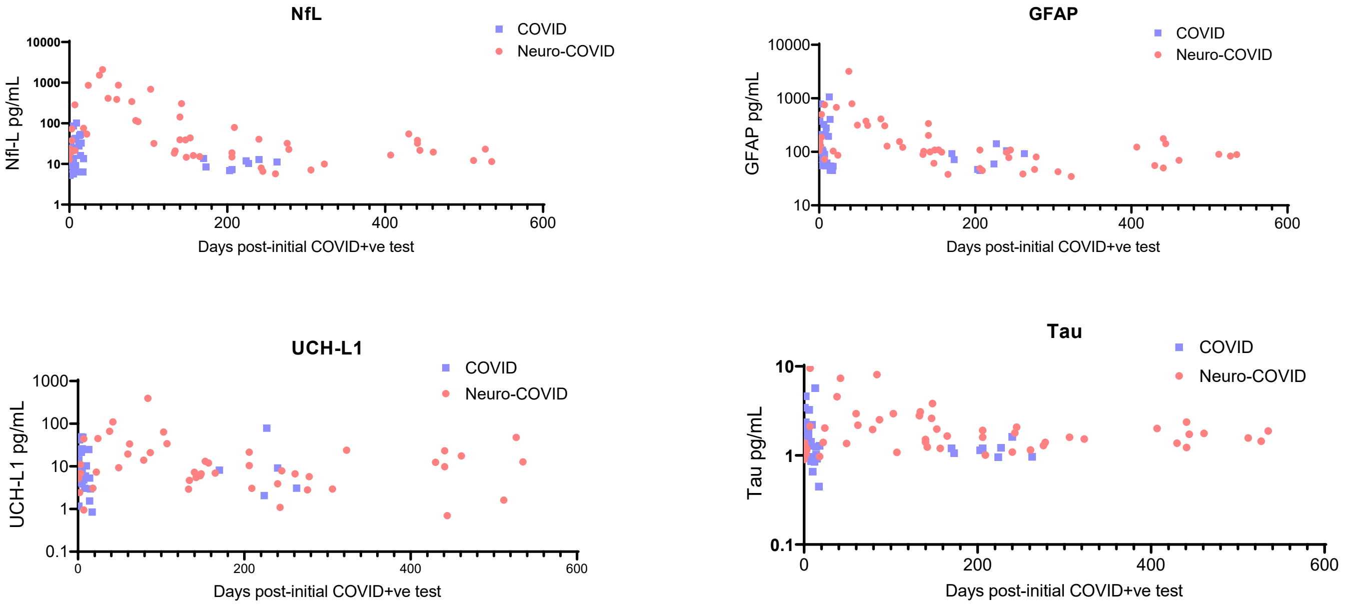


**Extended Data Fig. 1: COVID-CNS Brain injury markers over time**
Serum NfL, GFAP, UCH-L1 and Tau were measured by Simoa and plotted over time since initial COVID+ve test in the COVID-CNS (sub-acute and convalescent participants)

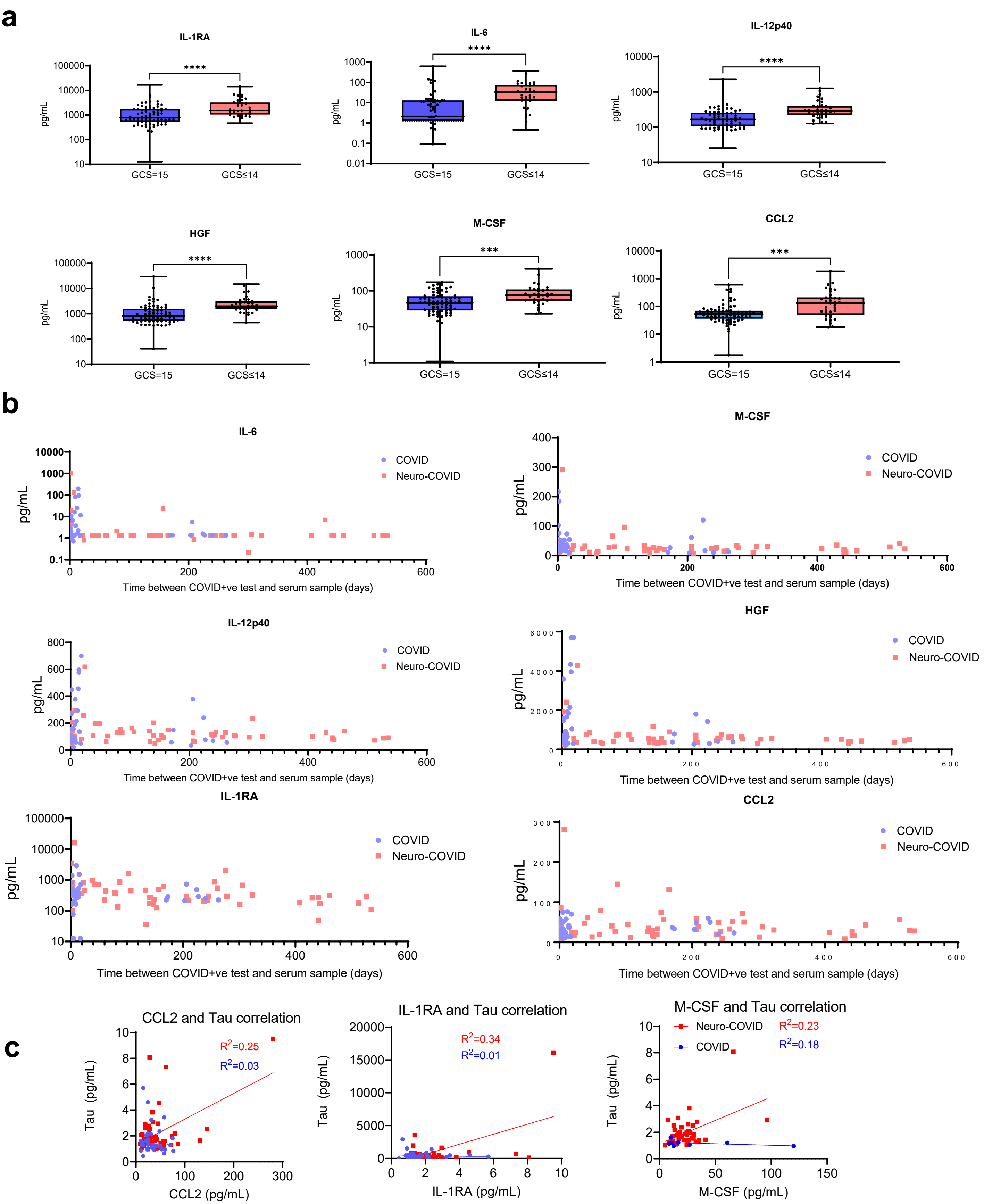


**Extended Data Fig. 2: Cytokine comparisons**

(a) Six cytokines measured by luminex were significantly higher in the sera of ISARIC participants (acute timepoints) with GCS≤14 compared to GCS=15 group (b) The six serum cytokines plotted over time since initial COVID+ve test in the COVID-CNS (sub-acute/convalescent) cohort (c) Significant correlations of three cytokines of interest with Tau in the COVID-CNS Neuro-COVID cohort (sub-acute/convalescent)

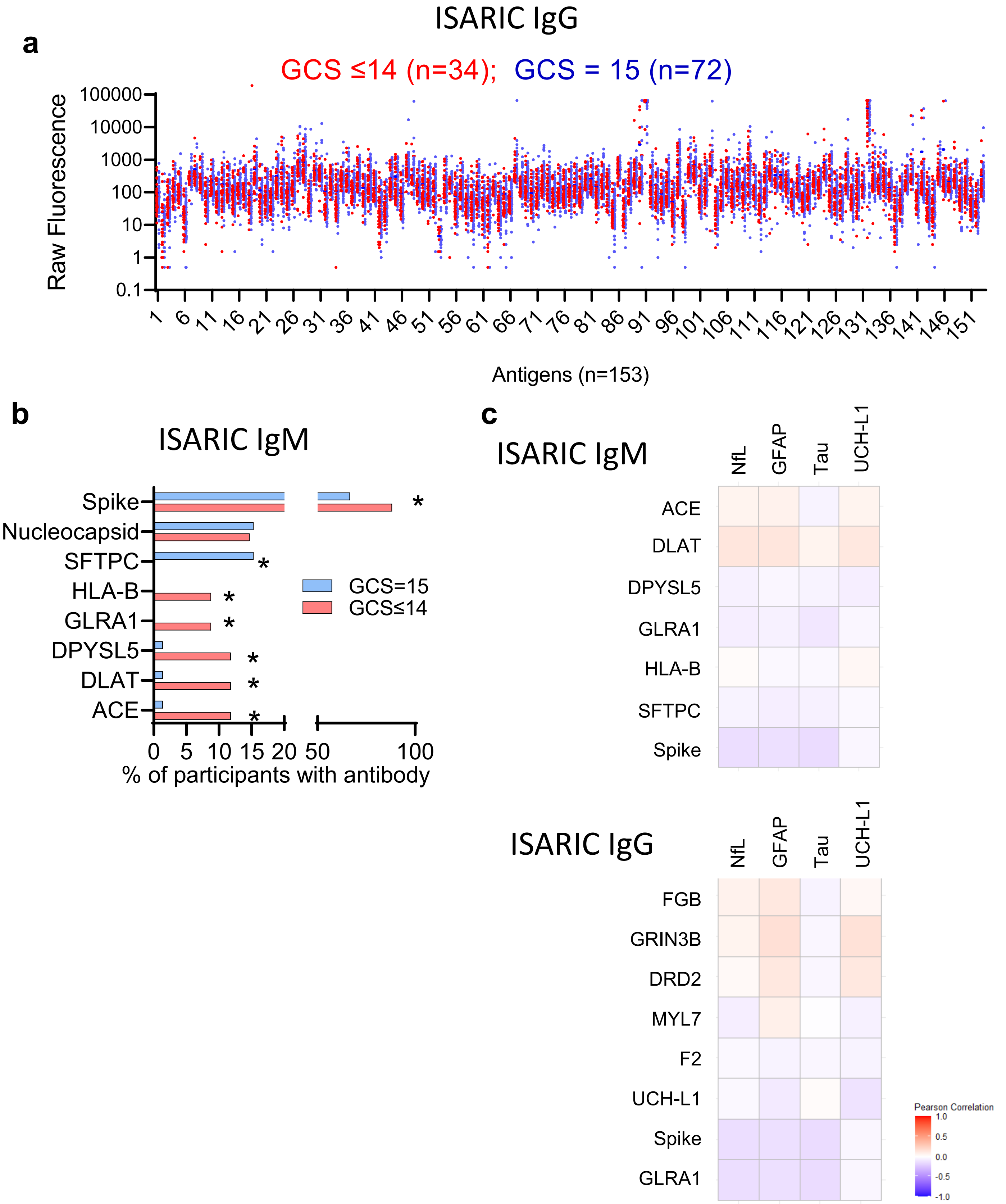


**Extended Data Fig. 3: ISARIC (acute) autoantibody screen**

(a) Raw fluorescence data from the protein chip used to screen antibodies against 153 antigens.

(b) Frequency of IgM autoantibodies in the ISARIC (acute) cohort (*Fisher's exact tests $p<0.05$)

(c) Heatmaps showing the insignificant correlations between IgM or IgG autoantibodies and brain injury markers, UCH-L1, Tau, GFAP, and NfL

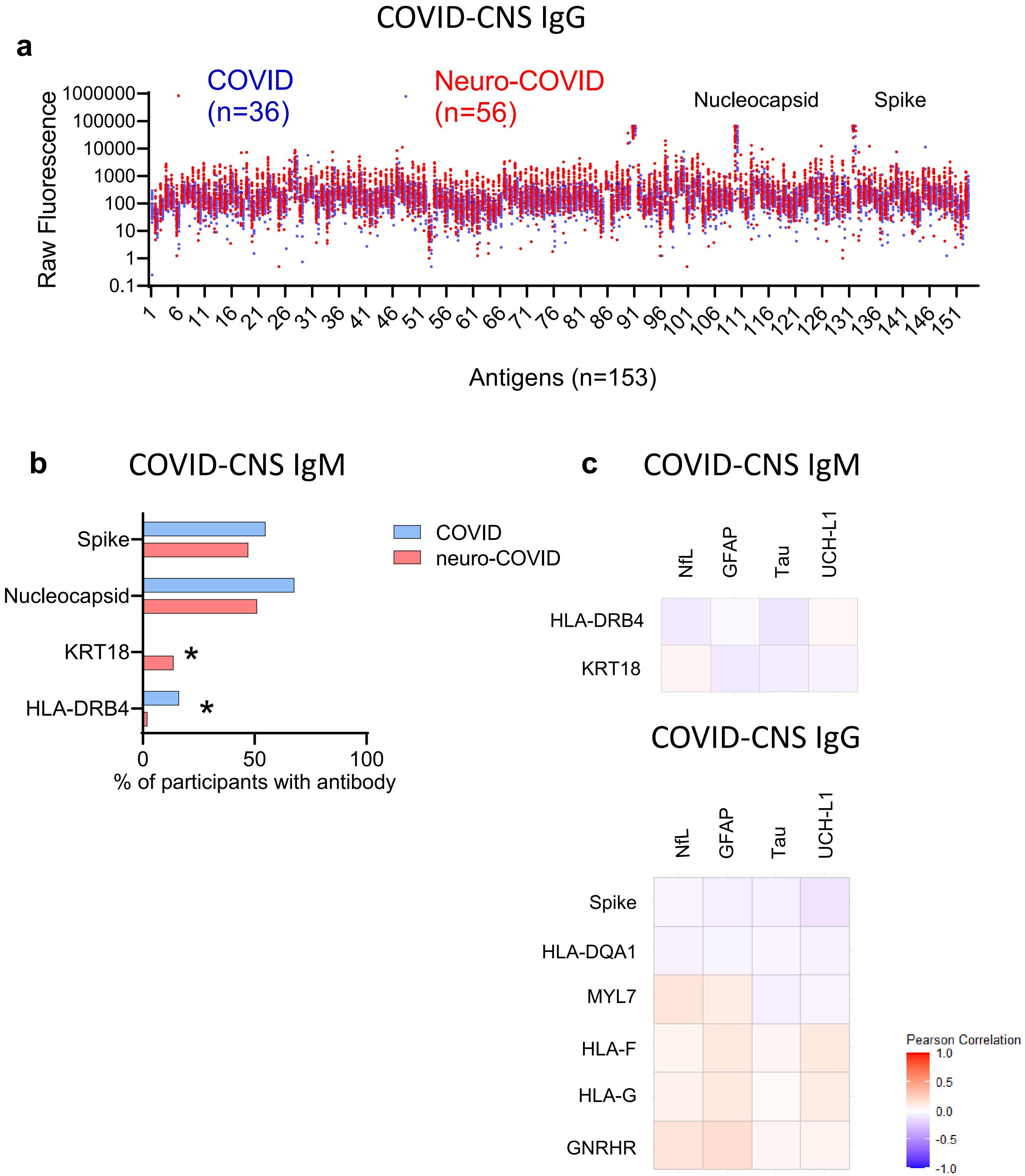


**Extended Data Fig 4: COVID-CNS (sub-acute/convalescent) autoantibody screen**

(a) Raw fluorescence data from the protein chip used to screen antibodies against 153 antigens

(b) Heatmaps showing the insignificant correlations between IgM or IgG autoantibodies and brain injury markers, UCH-L1, Tau, GFAP, and NfL

(c) Frequency of IgM autoantibodies in the COVID-CNS (sub-acute/convalescent) cohort (*Fisher's exact tests $p<0.05$)

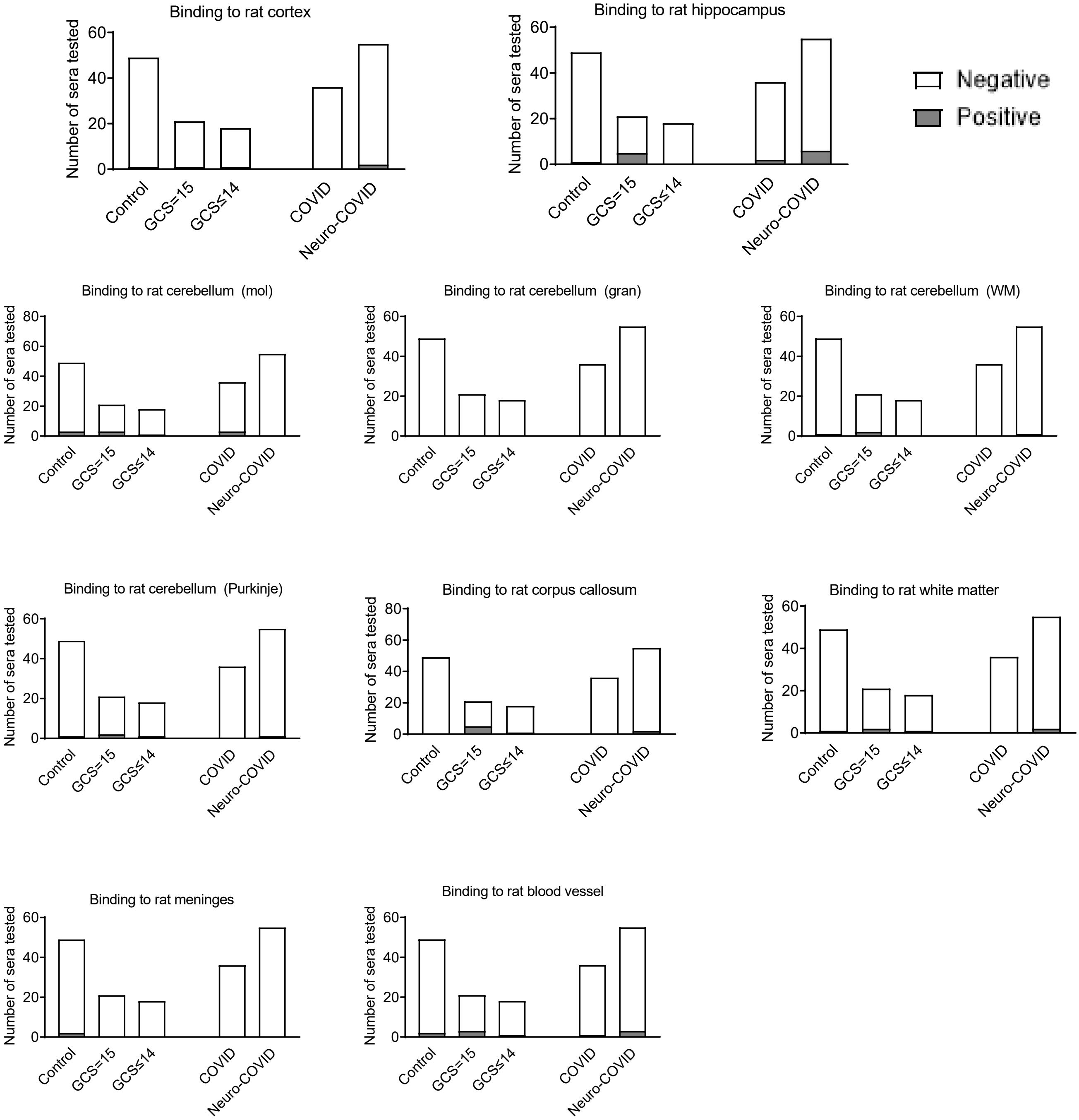


**Extended Data Fig. 5: CNS antibody screen**

Rat brain reactivity as detected by IHC in sera from ISARIC and COVID-CNS cohorts. The numbers positive were analysed by Fisher’s exact test with Benjamini and Hochberg correction. Only binding to brainstem showed significant differences between groups, as shown in main text Fig. 3. .

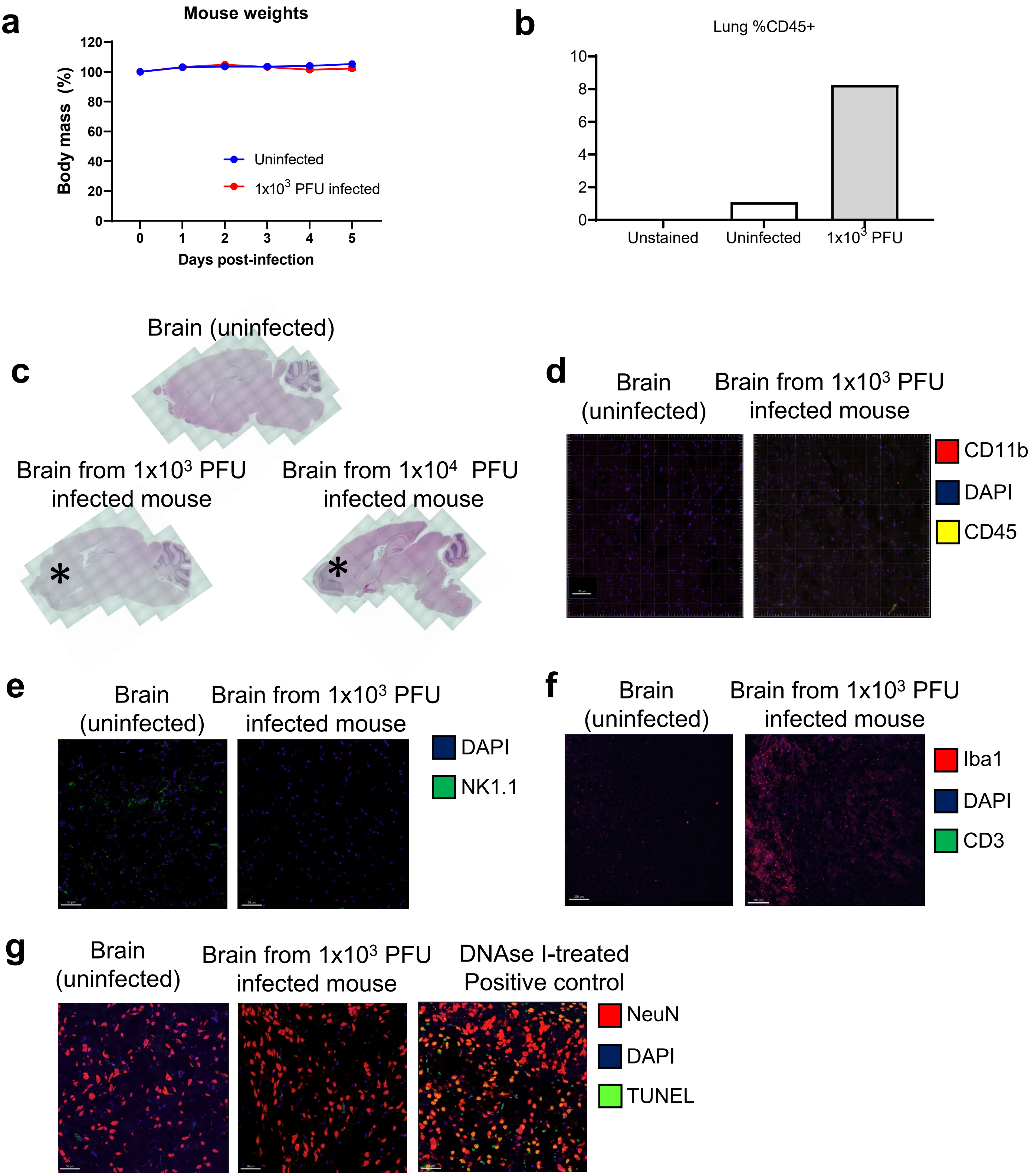


**Extended data Fig. 6: Mouse model of low-inoculum SARS-CoV-2 infection examined at day 5**

(a) Mouse weights over time

(b) Quantification of CD45 stain in mouse brain.

(c) H&E sagittal brain sections checked for mononuclear clusters

Confocal microscopy used to check for immune cells with different immunofluorescent stains:

(d) CD45 and CD11b

(e) NK1.1 and (f) CD3.

(g) Neuron viability was assayed my immunofluorescent staining and confocal microscopy

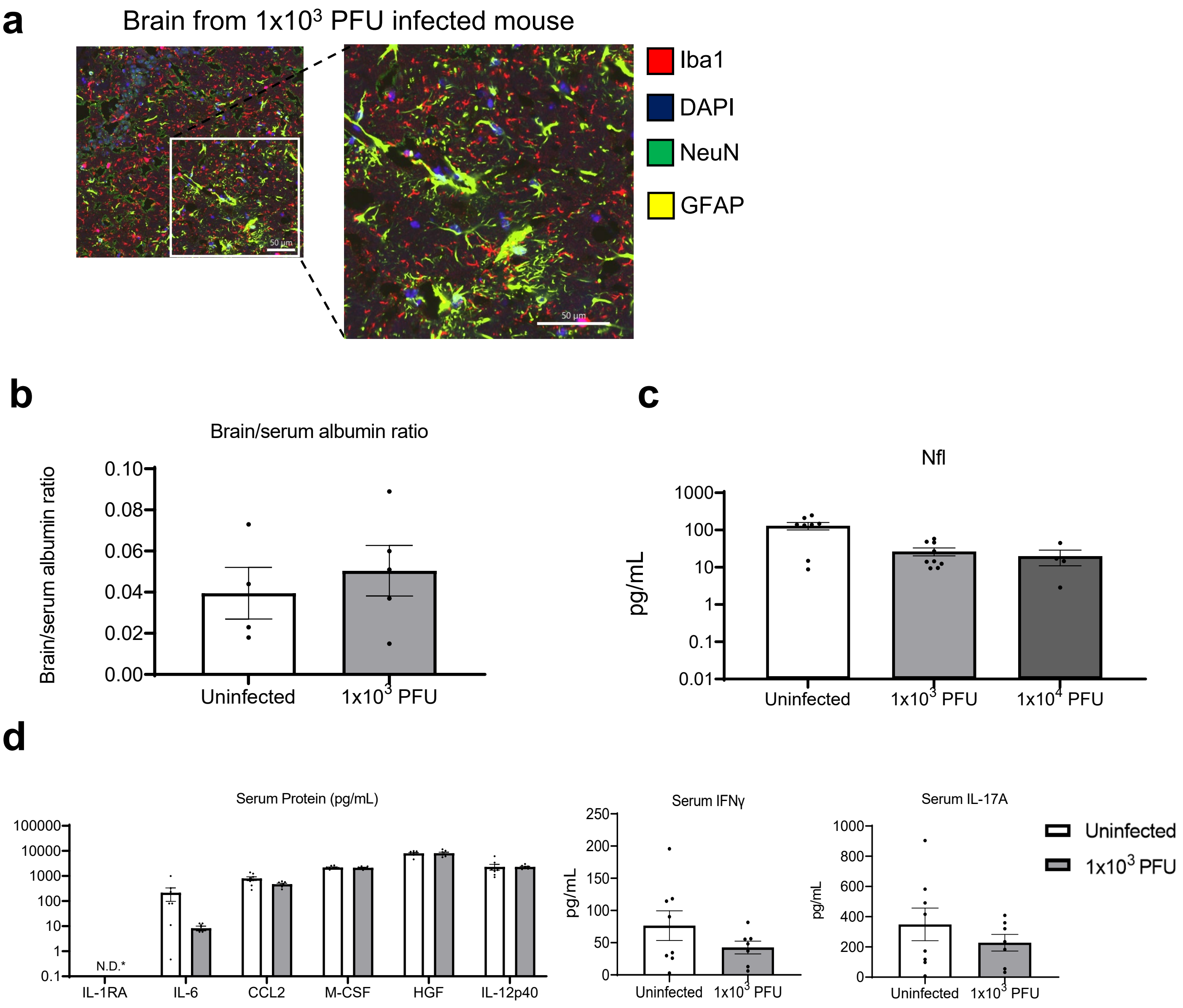


**Extended data Fig. 7: Brain and serum assays from mice infected with low-inoculum SARS-CoV-2 at day 5 post-infection**

(a) Clusters of GFAP+ microglia were observed co-localized with areas of high Iba1 density.

(b) Ratio of albumin in the brain and serum to assay blood brain barrier integrity determined by ELISA

(c) NfL measured in the serum by Simoa

(d) No significant differences in proteins were detected in mouse sera by Luminex *IL-RA undetectable by ELISA in all, but one uninfected serum sample (=3.5 pg/mL)

### Extended data Table 1: Median values of biomarkers

| Median (pg/mL) and (Q1, Q3) | NfL | GFAP | Tau | UCH-L1 | IL-1RA | IL-6 | IL-12p40 | CCL2 | M-CSF | HGF |
|---|---|---|---|---|---|---|---|---|---|---|
| **ISARIC GCS=15** | 26.5 (12.9, 62.3) | 125 (86.9, 230.0) | 1.1 (0.9, 1.4) | 21.9 (11.3, 36.3) | 772.5 (534.8, 1722.3) | 2.12 (1.4, 12.8) | 166.5 (109.3, 253.8) | 53.3 (36.6, 69.6) | 46.6 (28.5, 67.1) | 809.1 (530.5, 1505.0) |
| **ISARIC GCS≤14** | 58.2 (30.7, 122.8) | 151 (90.5, 263.0) | 1.2 (0.9, 1.7) | 33.7 (18.8, 48.5) | 1501.3 (1093.0, 3128.8) | 33.8 (12.6, 72.6) | 286.1 (224.1, 395.2) | 132.8 (55.5, 208.4) | 75.9 (54.0, 107.1) | 1965.0 (1570.0, 2956.6) |
| **COVID-CNS COVID** | 12.6 (8.3, 27.9) | 92.9 (61.3, 197.0) | 1.3 (1.1, 1.9) | 7.4 (4.0, 19.6) | 290.0 (197.2, 476.0) | 1.4 (1.4, 5.8) | 123.7 (68.4, 268.9) | 32.10 (23.1, 49.2) | 29.1 (17.6, 51.6) | 668.0 (385.9, 1693.2) |
| **COVID-CNS Neuro-COVID** | 32.3 (17.9, 111.8) | 106.5 (79.5, 191.0) | 1.7 (1.3, 2.2) | 9.3 (5.3, 20.9) | 303.1 (218.1, 546.7) | 1.36 (1.36, 1.36) | 108.8 (91.1, 141.9) | 32.5 (20.0, 50.0) | 24.4 (16.0, 30.5) | 529.9 (405.2, 727.6) |

### Extended Data Table 2: COVID-CNS Neuro-COVID cases

| | Condition, n (%) | Diagnosis | n |
|---|---|---|---|
| **Central Nervous System Condition 41 (73%)** | Cerebrovascular 12 (21%) | Ischaemic stroke | 6 |
| | | Haemorrhagic stroke | 2 |
| | | Cerebral venous sinus thrombosis | 1 |
| | | Vasculitic stroke | 1 |
| | | CNS vasculitis | 1 |
| | | PRES* | 1 |
| | CNS inflammation 14 (25%) | Encephalopathy | 5 |
| | | Encephalitis | 4 |
| | | Demyelinating | 5 |
| | Movement disorder 3 (5%) | Hyperkinetic movement disorder | 3 |
| | Seizures and others 12 (21%) | Seizures | 8 |
| | | Neuropsychiatric | 2 |
| | | Others | 2 |
| **Peripheral Nervous System Condition 15 (27%)** | Various PNS 15 (27%) | Peripheral nerves dysfunction | 11 |
| | | Myopathy | 2 |
| | | Cranial nerve palsy | 2 |

*PRES = posterior reversible encephalopathy syndrome

### Extended Data Table 3: ISARIC cohort cytokines correlations With Brain injury markers

| | Variables | NFL | | GFAP | | Tau | | UCHL | |
|---|---|---|---|---|---|---|---|---|---|
| | | Correlation | Adj p-value (Estimate) | Correlation | Adj p-value (Estimate) | Correlation | Adj p-value (Estimate) | Correlation | Adj p-value (Estimate) |
| **1** | **CCL5** | -0.168 | 1.000 | -0.190 | 1.000 | -0.157 | 1.000 | -0.023 | 1.000 |
| **2** | **SCGF-beta** | -0.086 | 1.000 | 0.018 | 1.000 | -0.020 | 1.000 | -0.012 | 1.000 |
| **3** | **CXCL10** | -0.056 | 1.000 | -0.086 | 1.000 | -0.031 | 1.000 | -0.042 | 1.000 |
| **4** | **IL-6** | -0.013 | 1.000 | -0.054 | 1.000 | -0.007 | 1.000 | 0.081 | 1.000 |
| **5** | **CCL2** | 0.118 | 1.000 | 0.106 | 1.000 | 0.098 | 1.000 | 0.070 | 1.000 |
| **6** | **CCL27** | 0.128 | 1.000 | 0.124 | 1.000 | 0.125 | 1.000 | 0.054 | 1.000 |
| **7** | **Eotaxin** | 0.091 | 1.000 | 0.056 | 1.000 | 0.161 | 1.000 | -0.019 | 1.000 |
| **8** | **CXCL9** | 0.120 | 1.000 | -0.067 | 1.000 | 0.075 | 1.000 | -0.004 | 1.000 |
| **9** | **IL-2Ra** | 0.067 | 1.000 | 0.019 | 1.000 | 0.064 | 1.000 | 0.114 | 1.000 |
| **10** | **HGF** | 0.616 | <0.001* (0.033) | 0.250 | 0.773 | 0.682 | <0.001* (0.0005) | 0.156 | 1.000 |
| **11** | **IL-12(p40)** | 0.632 | <0.001* (0.414) | 0.401 | 0.002* (1.027) | 0.651 | <0.001* (0.005) | 0.367 | 0.004* (0.096) |
| **12** | **IL-10** | 0.180 | 1.000 | 0.399 | 0.004* (30.74) | 0.119 | 1.000 | 0.610 | <0.001* (4.932) |
| **13** | **TNF** | 0.239 | 0.464 | 0.460 | <0.001* (9.518) | 0.167 | 1.000 | 0.561 | <0.001* (1.246) |
| **14** | **IL-2** | 0.222 | 1.000 | 0.534 | <0.001* (42.03) | 0.112 | 1.000 | 0.764 | <0.001* (6.439) |
| **15** | **IL-3** | 0.212 | 1.000 | 0.567 | <0.001* (965.6) | 0.140 | 1.000 | 0.758 | <0.001* (137.331) |
| **16** | **LTalpha** | -0.077 | 1.000 | 0.204 | 1.000 | -0.078 | 1.000 | 0.386 | 0.002* (0.184) |
| **17** | **IL-9** | 0.003 | 1.000 | 0.266 | 0.288 | 0.030 | 1.000 | 0.487 | <0.001* (0.307) |
| **18** | **IL-12(p70)** | 0.093 | 1.000 | 0.330 | 0.091 | 0.095 | 1.000 | 0.379 | 0.004* (1.807) |
| **19** | **IL-13** | -0.040 | 1.000 | 0.047 | 1.000 | 0.013 | 1.000 | 0.052 | 1.000 |
| **20** | **GM-CSF** | 0.060 | 1.000 | 0.110 | 1.000 | 0.046 | 1.000 | 0.154 | 1.000 |
| **21** | **IL-5** | 0.065 | 1.000 | 0.154 | 1.000 | 0.045 | 1.000 | 0.299 | 0.116 |
| **22** | **b-NGF** | 0.041 | 1.000 | 0.180 | 1.000 | 0.007 | 1.000 | 0.407 | 0.001* (5.598) |
| **23** | **CCL4** | -0.009 | 1.000 | -0.007 | 1.000 | -0.051 | 1.000 | 0.347 | 0.015* (0.115) |
| **24** | **IL-15** | 0.003 | 1.000 | -0.035 | 1.000 | -0.050 | 1.000 | 0.273 | 0.258 |
| **25** | **IL-8** | 0.010 | 1.000 | -0.025 | 1.000 | -0.028 | 1.000 | 0.297 | 0.109 |
| **26** | **CCL3** | -0.003 | 1.000 | -0.034 | 1.000 | -0.043 | 1.000 | 0.282 | 0.193 |
| **27** | **G-CSF** | -0.023 | 1.000 | -0.019 | 1.000 | -0.030 | 1.000 | 0.296 | 0.120 |
| **28** | **VEGF** | 0.013 | 1.000 | -0.055 | 1.000 | -0.077 | 1.000 | 0.187 | 1.000 |
| **29** | **CXCL1** | -0.049 | 1.000 | 0.006 | 1.000 | -0.091 | 1.000 | 0.205 | 1.000 |
| **30** | **TRAIL** | 0.028 | 1.000 | 0.264 | 0.498 | 0.046 | 1.000 | 0.441 | <0.001* (4.545) |
| **31** | **IL-1alpha** | 0.227 | 1.000 | 0.306 | 0.299 | 0.144 | 1.000 | 0.528 | <0.001* (1.717) |
| **32** | **FGF basic** | 0.305 | 0.118 | 0.278 | 0.468 | 0.279 | 0.241 | 0.408 | 0.001* (1.032) |
| **33** | **IFN-alpha2** | 0.173 | 1.000 | 0.006 | 1.000 | 0.178 | 1.000 | 0.296 | 0.110 |
| **34** | **IL-4** | 0.191 | 1.000 | 0.119 | 1.000 | 0.195 | 1.000 | 0.248 | 0.673 |
| **35** | **IL-17A** | 0.244 | 1.000 | 0.150 | 1.000 | 0.242 | 1.000 | 0.303 | 0.109 |
| **36** | **LIF** | 0.119 | 1.000 | 0.044 | 1.000 | 0.073 | 1.000 | 0.273 | 0.316 |
| **37** | **MIF** | 0.037 | 1.000 | -0.056 | 1.000 | -0.008 | 1.000 | 0.191 | 1.000 |
| **38** | **IL-1Ra** | 0.188 | 1.000 | 0.017 | 1.000 | 0.079 | 1.000 | 0.247 | 0.572 |
| **39** | **CCL7** | 0.098 | 1.000 | 0.045 | 1.000 | 0.022 | 1.000 | 0.140 | 1.000 |
| **40** | **IFN-gamma** | 0.194 | 1.000 | 0.169 | 1.000 | 0.163 | 1.000 | 0.276 | 0.157 |
| **41** | **SCF** | 0.334 | 0.007* (0.419) | 0.113 | 0.889 | 0.304 | 0.009* (0.005) | 0.237 | 0.213 |
| **42** | **M-CSF** | 0.293 | 0.094 | 0.112 | 1.000 | 0.211 | 0.727 | 0.234 | 0.359 |
| **43** | **IL-16** | 0.206 | 1.000 | 0.117 | 1.000 | 0.156 | 1.000 | 0.148 | 1.000 |
| **44** | **IL-18** | 0.205 | 1.000 | 0.036 | 1.000 | 0.113 | 1.000 | 0.173 | 1.000 |
| **45** | **IL-1beta** | 0.195 | 1.000 | 0.112 | 1.000 | 0.160 | 1.000 | 0.328 | 0.037* (7.381) |
| **46** | **PDGF-bb** | -0.065 | 1.000 | -0.125 | 1.000 | -0.163 | 1.000 | 0.114 | 1.000 |
| **47** | **CXCL12** | 0.081 | 1.000 | 0.140 | 1.000 | 0.118 | 1.000 | 0.271 | 0.375 |
| **48** | **IL-7** | -0.001 | 1.000 | 0.024 | 1.000 | -0.030 | 1.000 | 0.121 | 1.000 |

### Extended Data Table 4: ISARIC Median-centred data Differences between GCS=15 and GCS≤14

| Count | Cytokines | Adj p-value[1] (test,p-value[2]) | Effect size |
|---|---|---|---|
| 1 | CCL5 | 1 (M, <0.001) | -0.018 |
| 2 | SCGF-beta | 1 (M, <0.001) | -0.042 |
| 3 | CXCL10 | 0.717 (M, <0.001) | -0.232 |
| 4 | IL-6 | <0.001* (M, <0.001) | -0.429 |
| 5 | CCL2 | 0.137 (M, <0.001) | -0.284 |
| 6 | CCL27 | 0.16 (M, <0.001) | 0.279 |
| 7 | Eotaxin | <0.001* (M, <0.001) | 0.436 |
| 8 | CXCL9 | 1 (M, <0.001) | -0.103 |
| 9 | IL-2Ra | 1 (M, <0.001) | -0.028 |
| 10 | HGF | <0.001* (M, <0.001) | -0.425 |
| 11 | IL-12(p40) | 0.034* (M, <0.001) | -0.322 |
| 12 | IL-10 | 1 (M, <0.001) | -0.156 |
| 13 | TNF | 1 (M, <0.001) | 0.04 |
| 14 | IL-2 | 1 (M, <0.001) | 0.116 |
| 15 | IL-3 | 0.022* (M, <0.001) | 0.333 |
| 16 | LTalpha | 0.168 (M, <0.001) | 0.277 |
| 17 | IL-9 | 0.003* (M, <0.001) | 0.383 |
| 18 | IL-12(p70) | 0.127 (M, <0.001) | 0.286 |
| 19 | IL-13 | 1 (M, <0.001) | 0.07 |
| 20 | GM-CSF | 1 (M, <0.001) | -0.02 |
| 21 | IL-5 | 0.019* (M, <0.001) | 0.336 |
| 22 | b-NGF | 1 (M, <0.001) | 0.216 |
| 23 | CCL4 | 0.002* (M, <0.001) | 0.388 |
| 24 | IL-15 | 0.125 (M, <0.001) | 0.286 |
| 25 | IL-8 | 0.453 (M, <0.001) | -0.247 |
| 26 | CCL3 | 1 (M, <0.001) | -0.103 |
| 27 | G-CSF | 1 (M, <0.001) | -0.011 |
| 28 | VEGF | 1 (M, <0.001) | 0.033 |
| 29 | CXCL1 | 1 (M, <0.001) | -0.066 |
| 30 | TRAIL | 0.014* (M, <0.001) | 0.345 |
| 31 | IL-1alpha | 1 (M, <0.001) | 0.012 |
| 32 | FGF basic | 0.018* (M, <0.001) | 0.339 |
| 33 | IFN-alpha2 | 1 (M, <0.001) | -0.093 |
| 34 | IL-4 | <0.001* (M, <0.001) | 0.46 |
| 35 | IL-17A | 1 (M, <0.001) | -0.022 |
| 36 | LIF | 1 (M, <0.001) | 0.154 |
| 37 | MIF | 1 (M, <0.001) | -0.066 |
| 38 | IL-1Ra | 0.071* (M, <0.001) | -0.302 |
| 39 | CCL7 | 1 (M, <0.001) | -0.092 |
| 40 | IFN-gamma | 1 (M, <0.001) | -0.135 |
| 41 | SCF | 1 (M, <0.001) | 0.185 |
| 42 | M-CSF | 0.611 (M, <0.001) | -0.236 |
| 43 | IL-16 | 1 (M, <0.001) | -0.145 |
| 44 | IL-18 | 1 (M, <0.001) | -0.204 |
| 45 | IL-1beta | 0.421 (M, <0.001) | 0.249 |
| 46 | PDGF-bb | 1 (M, <0.001) | -0.146 |
| 47 | CXCL12 | 1 (M, <0.001) | 0.102 (M, <0.001) |
| 48 | IL-7 | 1 (M, <0.001) | 0.001 (M, <0.001) |

### Extended Data Table 5: Correlations of cytokines and brain injury markers in the COVID-CNS Neuro-COVID cohort with regression models

| Variables | NFL | | GFAP | | Tau | | UCHL | |
|---|---|---|---|---|---|---|---|---|
| | Correlation | Adj p-value | Correlation | Adj p-value | Correlation | Adj p-value | Correlation | Adj p-value |
| **CCL5** | -0.245 | 1.000 | -0.222 | 1.000 | 0.158 | 1.000 | 0.128 | 1.000 |
| **SCGF-beta** | 0.169 | 1.000 | 0.283 | 1.000 | 0.256 | 1.000 | 0.100 | 1.000 |
| **CXCL10** | 0.096 | 1.000 | 0.078 | 1.000 | 0.416 | 0.140 | 0.069 | 1.000 |
| **IL-6** | -0.059 | 1.000 | -0.010 | 1.000 | 0.003 | 1.000 | -0.043 | 1.000 |
| **CCL2** | 0.083 | 1.000 | 0.128 | 1.000 | 0.492 | 0.016* | 0.022 | 1.000 |
| **CCL27** | 0.183 | 1.000 | 0.032 | 1.000 | 0.255 | 1.000 | 0.006 | 1.000 |
| **Eotaxin** | -0.042 | 1.000 | 0.010 | 1.000 | -0.077 | 1.000 | -0.100 | 1.000 |
| **CXCL9** | 0.017 | 1.000 | -0.059 | 1.000 | 0.132 | 1.000 | 0.159 | 1.000 |
| **IL-2Ra** | 0.104 | 1.000 | 0.049 | 1.000 | 0.547 | 0.002* | 0.517 | 0.007* |
| **HGF** | 0.251 | 1.000 | 0.162 | 1.000 | 0.238 | 1.000 | 0.084 | 1.000 |
| **IL-12(p40)** | 0.298 | 1.000 | 0.116 | 1.000 | 0.200 | 1.000 | 0.124 | 1.000 |
| **IL-10** | -0.068 | 1.000 | -0.004 | 1.000 | 0.151 | 1.000 | -0.032 | 1.000 |
| **TNF** | 0.064 | 1.000 | 0.167 | 1.000 | 0.409 | 0.165 | 0.042 | 1.000 |
| **IL-2** | 0.064 | 1.000 | 0.111 | 1.000 | 0.123 | 1.000 | -0.129 | 1.000 |
| **IL-3** | NA | NA | NA | NA | NA | NA | NA | NA |
| **LTalpha** | 0.091 | 1.000 | 0.161 | 1.000 | -0.069 | 1.000 | -0.071 | 1.000 |
| **IL-9** | -0.009 | 1.000 | 0.075 | 1.000 | -0.051 | 1.000 | -0.082 | 1.000 |
| **IL-12(p70)** | 0.043 | 1.000 | -0.123 | 1.000 | -0.139 | 1.000 | -0.028 | 1.000 |
| **IL-13** | -0.008 | 1.000 | -0.079 | 1.000 | -0.159 | 1.000 | -0.049 | 1.000 |
| **GM-CSF** | -0.079 | 1.000 | -0.091 | 1.000 | -0.161 | 1.000 | -0.038 | 1.000 |
| **IL-5** | -0.068 | 1.000 | -0.063 | 1.000 | -0.053 | 1.000 | -0.054 | 1.000 |
| **b-NGF** | -0.078 | 1.000 | -0.070 | 1.000 | -0.044 | 1.000 | -0.042 | 1.000 |
| **CCL4** | 0.143 | 1.000 | 0.176 | 1.000 | -0.044 | 1.000 | -0.036 | 1.000 |
| **IL-15** | -0.031 | 1.000 | -0.016 | 1.000 | -0.080 | 1.000 | -0.009 | 1.000 |
| **IL-8** | -0.172 | 1.000 | -0.118 | 1.000 | 0.246 | 1.000 | 0.152 | 1.000 |
| **CCL3** | 0.094 | 1.000 | 0.052 | 1.000 | 0.006 | 1.000 | 0.008 | 1.000 |
| **G-CSF** | 0.086 | 1.000 | 0.044 | 1.000 | 0.031 | 1.000 | 0.001 | 1.000 |
| **VEGF** | -0.078 | 1.000 | -0.071 | 1.000 | -0.009 | 1.000 | -0.039 | 1.000 |
| **CXCL1** | -0.060 | 1.000 | -0.067 | 1.000 | -0.171 | 1.000 | -0.050 | 1.000 |
| **TRAIL** | 0.009 | 1.000 | -0.022 | 1.000 | 0.167 | 1.000 | -0.080 | 1.000 |
| **IL-1alpha** | -0.006 | 1.000 | -0.012 | 1.000 | 0.069 | 1.000 | 0.043 | 1.000 |
| **FGF basic** | 0.259 | 1.000 | 0.189 | 1.000 | 0.251 | 1.000 | 0.141 | 1.000 |
| **IFN-alpha2** | 0.034 | 1.000 | 0.052 | 1.000 | 0.368 | 0.432 | -0.003 | 1.000 |
| **IL-4** | -0.006 | 1.000 | 0.034 | 1.000 | -0.009 | 1.000 | -0.015 | 1.000 |
| **IL-17A** | -0.074 | 1.000 | 0.004 | 1.000 | 0.002 | 1.000 | -0.108 | 1.000 |
| **LIF** | -0.100 | 1.000 | -0.087 | 1.000 | 0.103 | 1.000 | 0.135 | 1.000 |
| **MIF** | 0.061 | 1.000 | 0.102 | 1.000 | 0.303 | 1.000 | 0.074 | 1.000 |
| **IL-1Ra** | 0.069 | 1.000 | 0.185 | 1.000 | 0.582 | 0.001* | 0.039 | 1.000 |
| **CCL7** | 0.033 | 1.000 | 0.158 | 1.000 | 0.608 | <0.001* | 0.050 | 1.000 |
| **IFN-gamma** | -0.035 | 1.000 | 0.019 | 1.000 | 0.177 | 1.000 | 0.003 | 1.000 |
| **SCF** | 0.039 | 1.000 | 0.119 | 1.000 | 0.621 | <0.001* | 0.084 | 1.000 |
| **M-CSF** | 0.077 | 1.000 | 0.139 | 1.000 | 0.680 | <0.001* | 0.231 | 1.000 |
| **IL-16** | 0.048 | 1.000 | 0.157 | 1.000 | 0.578 | 0.001* | 0.098 | 1.000 |
| **IL-18** | 0.132 | 1.000 | 0.190 | 1.000 | 0.724 | <0.001* | 0.317 | 1.000 |
| **IL-1beta** | 0.100 | 1.000 | -0.007 | 1.000 | 0.337 | 0.844 | 0.072 | 1.000 |
| **PDGF-bb** | 0.181 | 1.000 | 0.186 | 1.000 | -0.005 | 1.000 | -0.058 | 1.000 |
| **CXCL12** | 0.107 | 1.000 | 0.201 | 1.000 | 0.091 | 1.000 | -0.091 | 1.000 |
| **IL-7** | -0.030 | 1.000 | -0.007 | 1.000 | -0.029 | 1.000 | -0.040 | 1.000 |

### Extended Data Table 6: List of antigens on protein array

| Number | Antigen | Type | Number | Antigen | Type | Number | Antigen | Type |
|---|---|---|---|---|---|---|---|---|
| 1 | ACE | Ubiquitous | 52 | GNRHR | Endocrine | 103 | MYL4 | Cardiac |
| 2 | ACTA1 | Muscle | 53 | GRIA2 | Central Nervous System | 104 | MYL7 | Cardiac |
| 3 | ADAMTS13 | Coagulation | 54 | GRIA3 | Central Nervous System | 105 | NEFL | Central Nervous System |
| 4 | AGER | Lung | 55 | GRIA4 | Central Nervous System | 106 | NOVA1 | Central Nervous System |
| 5 | AGTR1 | Kidney | 56 | GRIN1 | Central Nervous System | 107 | NPHS2 | Kidney |
| 6 | ANKRD23 | Muscle | 57 | GRIN2A | Central Nervous System | 108 | NPPA | Cardiac |
| 7 | ANXA4 | Central Nervous System | 58 | GRIN3A | Central Nervous System | 109 | NPPB | Cardiac |
| 8 | ANXA5 | Coagulation | 59 | GRIN3B | Central Nervous System | 110 | Nucleocapsid protein | SARS-CoV-2 |
| 9 | APOH | Coagulation | 60 | GRINA | Central Nervous System | 111 | NUP210 | Ubiquitous |
| 10 | APP | Central Nervous System | 61 | GRM1 | Central Nervous System | 112 | OMG | Central Nervous System |
| 11 | BSG | Central Nervous System | 62 | GRM2 | Central Nervous System | 113 | PLA2R1 | Kidney |
| 12 | CD74 | HLA | 63 | GRM3 | Central Nervous System | 114 | PNMA1 | Central Nervous System |
| 13 | CDH1 | Ubiquitous | 64 | GRM4 | Central Nervous System | 115 | PNMA2 | Central Nervous System |
| 14 | CDH13 | Central Nervous System | 65 | GRM7 | Central Nervous System | 116 | POMC | Endocrine |
| 15 | CDR2 | Central Nervous System | 66 | GRM8 | Central Nervous System | 117 | PPP1R27 | Muscle |
| 16 | CEACAM1 | Ubiquitous | 67 | GSTT1 | Kidney | 118 | PRL | Endocrine |
| 17 | CEACAM5 | Ubiquitous | 68 | HARS | Muscle | 119 | PROC | Coagulation |
| 18 | CENPB | Ubiquitous | 69 | HLA-A | HLA | 120 | PROS1 | Coagulation |
| 19 | CENPH | Ubiquitous | 70 | HLA-B | HLA | 121 | PRTN3 | Kidney |
| 20 | CHRM2 | Cardiac | 71 | HLA-C | HLA | 122 | S100B | Central Nervous System |
| 21 | CHRNA10 | Muscle | 72 | HLA-DMA | HLA | 123 | SCGB1A1 | Lung |
| 22 | CHRNA9 | Central Nervous System | 73 | HLA-DMB | HLA | 124 | SCGB3A2 | Lung |
| 23 | CLDN5 | BBB | 74 | HLA-DOA | HLA | 125 | SELE | BBB |
| 24 | COL1A1 | Lung | 75 | HLA-DOB | HLA | 126 | SFTPA1 | Lung |
| 25 | COL1A2 | Lung | 76 | HLA-DPA1 | HLA | 127 | SFTPA2 | Lung |
| 26 | COL4a3 | Kidney | 77 | HLA-DPB1 | HLA | 128 | SFTPC | Lung |
| 27 | COL4A3BP | Central Nervous System | 78 | HLA-DQA1 | HLA | 129 | SLC22A12 | Kidney |
| 28 | COL5A2 | Lung | 79 | HLA-DQB1 | HLA | 130 | SLC2A1 | BBB |
| 29 | DBT | Ubiquitous | 80 | HLA-DQB2 | HLA | 131 | SNCA | Central Nervous System |
| 30 | DCN | Central Nervous System | 81 | HLA-DRA | HLA | 132 | Spike protein | SARS-CoV-2 |
| 31 | DDC | Ubiquitous | 82 | HLA-DRB1 | HLA | 133 | SSB | Central Nervous System |
| 32 | DLAT | Ubiquitous | 83 | HLA-DRB3 | HLA | 134 | TGM2 | Ubiquitous |
| 33 | DPYSL5 | Central Nervous System | 84 | HLA-DRB4 | HLA | 135 | TJP1 | BBB |
| 34 | DRD2 | Central Nervous System | 85 | HLA-DRB5 | HLA | 136 | TMEM174 | Kidney |
| 35 | DUPD1 | Muscle | 86 | HLA-E | HLA | 137 | TNNI3 | Cardiac |
| 36 | EDNRA | Cardiac | 87 | HLA-F | HLA | 138 | TNNT2 | Cardiac |
| 37 | ELAVL4 | Central Nervous System | 88 | HLA-G | HLA | 139 | TPH1 | Central Nervous System |
| 38 | F2 | Coagulation | 89 | IDI2 | Muscle | 140 | TPM1 | Muscle |
| 39 | F7 | Coagulation | 90 | IFNA1 | Ubiquitous | 141 | TPO | Endocrine |
| 40 | F8 | Coagulation | 91 | IGHG1 | Immune system | 142 | TROVE2 | Central Nervous System |
| 41 | F9 | Coagulation | 92 | KCNJ10 | Central Nervous System | 143 | TSHB | Endocrine |
| 42 | FGB | Coagulation | 93 | KRT18 | Ubiquitous | 144 | TSHR | Endocrine |
| 43 | FSHB | Endocrine | 94 | LAMC2 | BBB | 145 | TTN | Muscle |
| 44 | GABBR1 | Central Nervous System | 95 | LGI1 | Central Nervous System | 146 | TUBA1B | Lung |
| 45 | GABRA1 | Central Nervous System | 96 | LRRC10 | Cardiac | 147 | TUBB3 | Central Nervous System |
| 46 | GABRB3 | Central Nervous System | 97 | MAG | Central Nervous System | 148 | UCHL1 | Central Nervous System |
| 47 | GAD1 | Central Nervous System | 98 | MAPT | Central Nervous System | 149 | UCP3 | Muscle |
| 48 | GAD2 | Central Nervous System | 99 | MBP | Central Nervous System | 150 | UMOD | Kidney |
| 49 | GFAP | Central Nervous System | 100 | MOG | Central Nervous System | 151 | VIM | Kidney |
| 50 | GHRHR | Endocrine | 101 | MPO | Kidney | 152 | ZIC4 | Central Nervous System |
| 51 | GLRA1 | Central Nervous System | 102 | MYBPHL | Cardiac | 153 | ZNF397 | Ubiquitous |
